# Artificial Intelligence Models for Predicting Molecular Pathway Activity in Spinal Cord Injury: A Systematic Review

**DOI:** 10.1101/2025.10.07.25337258

**Authors:** Mahdi Mehmandoost, Mahtab Jabbar, Behnaz Rahatijafarabad, Mandana Mehrdad, Ibrahim Mohammadzadeh, Roozbeh Tavanaei, Sayeh Oveisi, Saeed Oraee-Yazdani, Alireza Zali, Farzan Fahim

## Abstract

**Background:** Spinal cord injury (SCI) remains a devastating neurological condition with high global incidence and minimal curative options. The pathobiology is multifactorial, encompassing acute mechanical insult, sustained neuroinflammation, oxidative stress, mitochondrial dysfunction, and glial scarring. Molecular signaling pathways orchestrate these responses, yet their clinical exploitation has been hindered by complex gene environment interactions and heterogeneous patient profiles. Artificial intelligence (AI) offers unprecedented analytical capacity to integrate multi-dimensional omics datasets, enabling prediction of pathway activities, identification of key biomarkers, and prioritization of therapeutic targets.

**Objectives:** To systematically synthesize current evidence on AI-driven prediction and modeling of molecular pathway processes in SCI, evaluate the performance and methodological quality of these approaches, and highlight translational opportunities for regenerative and precision medicine.

**Methods:** Following PRISMA guidelines, we searched PubMed, Scopus, and Web of Science (2000-2025) and registered the protocol in PROSPERO (CRD420251115723). Inclusion criteria encompassed studies applying AI algorithms to predict or model molecular pathway activity in SCI. Risk of bias was assessed with the PROBAST tool. Data on study design, cohort characteristics, AI methodology, pathway findings, biomarker signatures, and validation approaches were extracted and synthesized.

**Results:** Of 86 records, 11 studies met the criteria. Most were observational, bioinformatics-driven investigations published after 2020, predominantly from China, with heavy reliance on the GSE151371 blood transcriptomic dataset and small validation cohorts. Diagnostic and severity classification: Six studies achieved AUC values ranging 0.79-1.00; recurrent biomarkers included FCER1G, NFATC2, S100A8, and IL2RB. Overfitting risk was high due to dataset reuse and limited external validation. Mechanistic insights: Seven studies converged on immune dysregulation (NF-κB, VEGF, JAK-STAT, Toll-like receptor), novel cell death modalities (PANoptosis, cuproptosis), and metabolic/autophagy disruptions (PINK1/SQSTM1). Therapeutic predictions: Five studies proposed interventions drug repurposing (Emricasan, Alaproclate, Imatinib), cuproptosis-targeted agents, ZnO nanoparticles, and mesenchymal stem-cell transplantation with all validations restricted to preclinical models. GRADE assessment rated evidence as very low to low certainty, primarily due to risk of bias, indirectness, and imprecision.

**Conclusions:** AI has begun mapping the intricate immune metabolic degenerative network underpinning SCI, revealing candidate biomarkers and therapeutic targets with potential regenerative relevance. However, current evidence is constrained by small, homogeneous datasets, preclinical bias, and lack of longitudinal human validation. Scaling to multicenter, multi-omics cohorts and advancing promising candidates into early-phase trials are essential next steps to realize AI translational promise in SCI management.

## Introduction

Spinal cord injury (SCI) is a debilitating neurological condition that results in severe and often permanent motor and sensory deficits, with significant personal and socioeconomic impact. The majority of SCI cases arise from traumatic events such as vehicular accidents, falls, or sports injuries, and the global burden remains high according to the 2022 Global Burden of Disease Study, there were approximately 0.9□million new SCI cases in 2019, with 20.6□million individuals living with the condition worldwide□[1,□2]. Beyond the immediate mechanical insult, secondary injury cascades characterized by neuroinflammation, oxidative stress, excitotoxicity, glial scar formation, and progressive axonal degeneration continue to exacerbate neural loss and impair endogenous repair mechanisms□[1–3]. Understanding and modulating these secondary processes is essential for improving recovery outcomes.

Molecular signaling pathways represent the fundamental regulatory networks governing cellular responses to SCI. Dysregulation in pro-inflammatory cascades (e.g., NF-κB, JAK-STAT, toll-like receptor signaling), angiogenic regulators (e.g., VEGF), cell death programs (e.g.,□cuproptosis, PANoptosis, autophagy), and metabolic checkpoints (e.g., mitochondrial bioenergetics) can severely influence the trajectory of tissue damage and recovery□[3,□8,□11,□15,□16]. Identifying the key molecular drivers of SCI pathology is crucial for discovering actionable biomarkers and formulating targeted therapies that go beyond symptom management to promote neuroprotection and regeneration.

Artificial intelligence (AI), particularly machine learning (ML) and deep learning, has become an increasingly powerful tool for leveraging high-dimensional omics datasets in SCI research. Recent bioinformatics-driven studies have applied algorithms such as support vector machines, artificial neural networks, and weighted gene co-expression network analysis to transcriptomic data most notably the extensively used GSE151371 dataset to classify injury severity, predict molecular pathway activity, and uncover recurrent biomarkers [7–12]. Multiple investigations have reported impressive predictive performance (AUC□0.79–1.00), with immune-related genes including *FCER1G*, *S100A8*, and *IL2RB* repeatedly emerging as critical nodes of injury-associated networks□[7–12,□17]. These insights open opportunities for precision diagnostics and the design of pathway-targeted interventions.

### Comparison with Previous Reviews

Existing reviews have addressed AI applications in SCI from different perspectives, but none have systematically synthesized findings on AI-driven prediction of molecular pathway activity. For example, Kim□et□al.□[28] and Dietz□et□al.□[29] summarized the use of ML models such as SVM and CNN for improving diagnosis, outcome prediction, and rehabilitation planning in acute SCI, focusing primarily on clinical measures and imaging biomarkers, with limited integration of molecular data. Martín-Noguerol□et□al.□[30] focused on AI-assisted imaging techniques for detecting traumatic and degenerative spinal pathologies, aiming to improve radiological efficiency, but without exploring molecular mechanisms. Covache□et□al.□[5] discussed advanced regenerative strategies integrating CRISPR technologies, single-cell omics, and AI-driven therapeutics in neural tissue repair, yet did not address diagnostic biomarker modeling from a systems biology perspective. Notably, none of these reviews has systematically addressed the prediction of immune-, metabolic-, and cell death–related pathways from omics datasets, or critically examined their translational potential in regenerative medicine. This gap is significant given the convergence of multi-omics technologies, AI-based analytical capabilities, and the urgent clinical need for post-injury neuroregenerative strategies.

### Aim of the Present Review

The present systematic review was conducted to fill this gap by (i) consolidating current evidence on AI and ML applications for predicting molecular pathway activity following SCI, (ii) summarizing the performance and methodology of these models, and (iii) identifying recurrent molecular signatures and mechanistic themes that can inform targeted therapeutic development within a regenerative medicine framework. By critically evaluating the strengths, limitations, and translational prospects of AI-bioinformatics approaches, this work aims to provide a comprehensive resource for guiding future research and accelerating the bench-to-bedside translation of pathway-specific interventions for SCI.

## Method

### Study design

This work is a systematic review conducted in accordance with the Preferred Reporting Items for Systematic Reviews and Meta-Analyses (PRISMA) 2020 guidelines. The protocol was prospectively registered in PROSPERO (CRD420251115723). Although a meta-analysis was planned in the protocol stage, it was not performed due to substantial heterogeneity in study designs, datasets, AI model architectures, and reported outcome metrics among the included studies. The PRISMA checklist is provided in ***Supplementary***□***1****a*.

### Search strategy

A comprehensive search was performed in PubMed, Scopus, and Web of Science for studies published between 2000 to 2025. No language restrictions were applied, moreover, To ensure comprehensive coverage, we also searched Google Scholar and manually checked the reference lists of included studies and relevant reviews for additional references. A complete search syntax is available in ***Supplementary***□***1b*;** the core search string was:

(“spinal cord injury” OR “SCI” OR “spinal cord trauma”)

AND (“molecular pathway” OR “signaling pathway” OR “gene expression” OR “transcriptomic” OR “omics”)

AND (“artificial intelligence” OR “machine learning” OR “deep learning” OR “bioinformatics” OR “predict”)

### Eligibility Criteria

Inclusion criteria:

- Primary research studies using AI or machine learning to predict or model molecular pathway activity in the context of SCI.
- Analyses based on omics data (e.g., transcriptomic, proteomic, epigenomic) from human or relevant animal models.
- Studies reporting at least one AI performance metric (e.g., AUC, accuracy).

Exclusion criteria:

- Studies assessing only clinical outcomes without molecular analyses.
- Stem cell intervention studies where AI was not applied to pathway prediction.
- Non-original research (reviews, editorials, conference abstracts without full datasets).
- Studies with no extractable outcome metrics or insufficient methodological detail.

### Screening Process

Following the removal of duplicates, two reviewers independently (BR and MJ) screened titles, abstracts of included studies and subsequently, two other reviewers assessed and investigate the full texts of final included articles and also, Disagreements at any stage were resolved by consultation with a third reviewer (FF). the excluded sheets were also provided based in the full text screen processes by these two reviewers ***(supplementary 3a and 4a)*.** At all stages of the screening process, disagreements were resolved through discussion until consensus was reached.

### Data Extraction

Data from each included study were extracted independently by two reviewers (MM and MJ) into a standardized Excel spreadsheet. Extracted variables included:

- Bibliographic details (author, year, country)
- Study design and population/sample source
- AI methodology (algorithm type, feature selection method, validation strategy)
- Dataset characteristics (e.g., GSE number, sample size, human/animal origin)
- Molecular pathway categories and key biomarkers
- Reported performance metrics: AUC, accuracy, precision, recall, F1-score, specificity, sensitivity, effect size

***(Supplementary 3b and 4b.)***

### Outcomes

The primary outcome was AI model performance for predicting molecular pathway activity in SCI, expressed as standard diagnostic metrics (AUC, accuracy, precision, recall, F1-score, specificity, sensitivity, effect size).

Secondary outcomes included the identification of recurrent pathways and biomarkers, methodological trends, and translational recommendations.

### Risk of Bias Assessment

Risk of bias was assessed using the PROBAST risk of bias tool for diagnostic accuracy studies. ***(Supplementary 5 and 6.)*** this approach ensured that each study was evaluated utilizing the most suitable criteria for its design. Two reviewers independently evaluated the risk of bias, and any disagreements were resolved through discussion or by consulting a third reviewer. **(*Figure 1*.)**

**Fig 1.**
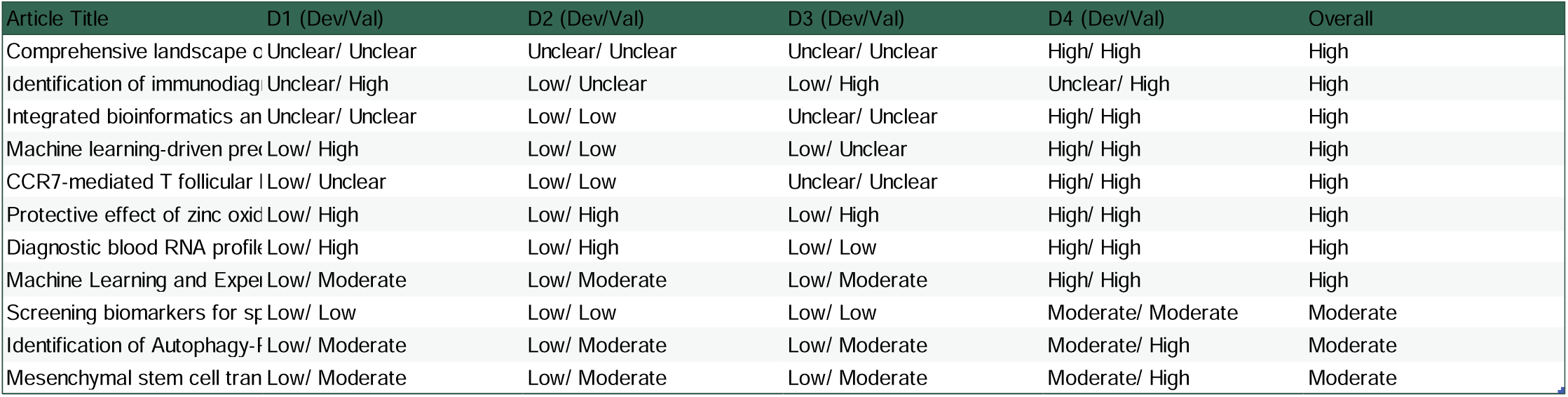
The risk of bias assessment of the study using PROBAST risk of bias tool.

### Results

The systematic literature search using the terms mentioned above yielded 86 records, comprising 19 studies identified in PubMed, 45 in Scopus, and 22 in Web of Science. After removing duplicates (8), 38 papers were discarded in the initial screening phase, since either their abstract or title didn’t match our inclusion criteria. The remaining 40 papers were compiled as full text and further assessed for eligibility. Among these, 11 records were included for further analyses in this systematic review according to the previously outlined criteria. At the same time, 30 articles were excluded as they met at least one of the exclusion criteria at the full-text level.

Our review included 11 studies ***(Table 1.)*** that shared notable similarities: all were published after 2020, most were from China (with one from the USA and another with UK collaborations), and almost all were observational, bioinformatics-driven studies with validation via animal models or qPCR. We also identified a heavy reliance on the GSE151371 transcriptomic dataset, with small sample sizes in both human and animal validation cohorts. **(*Figure 2*.)**

**Fig 2.**
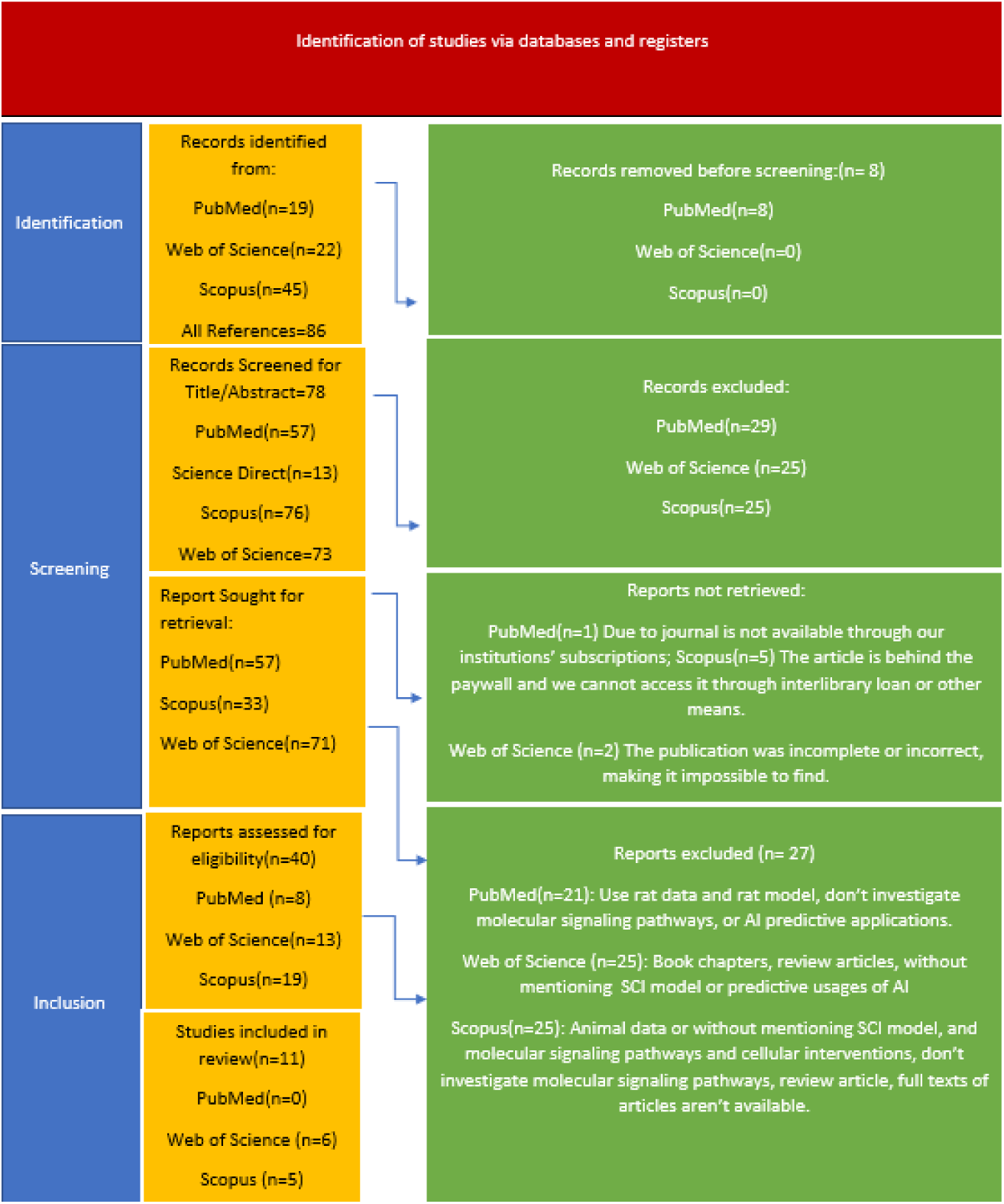
PRISMA flow chart.

**Table 1.**
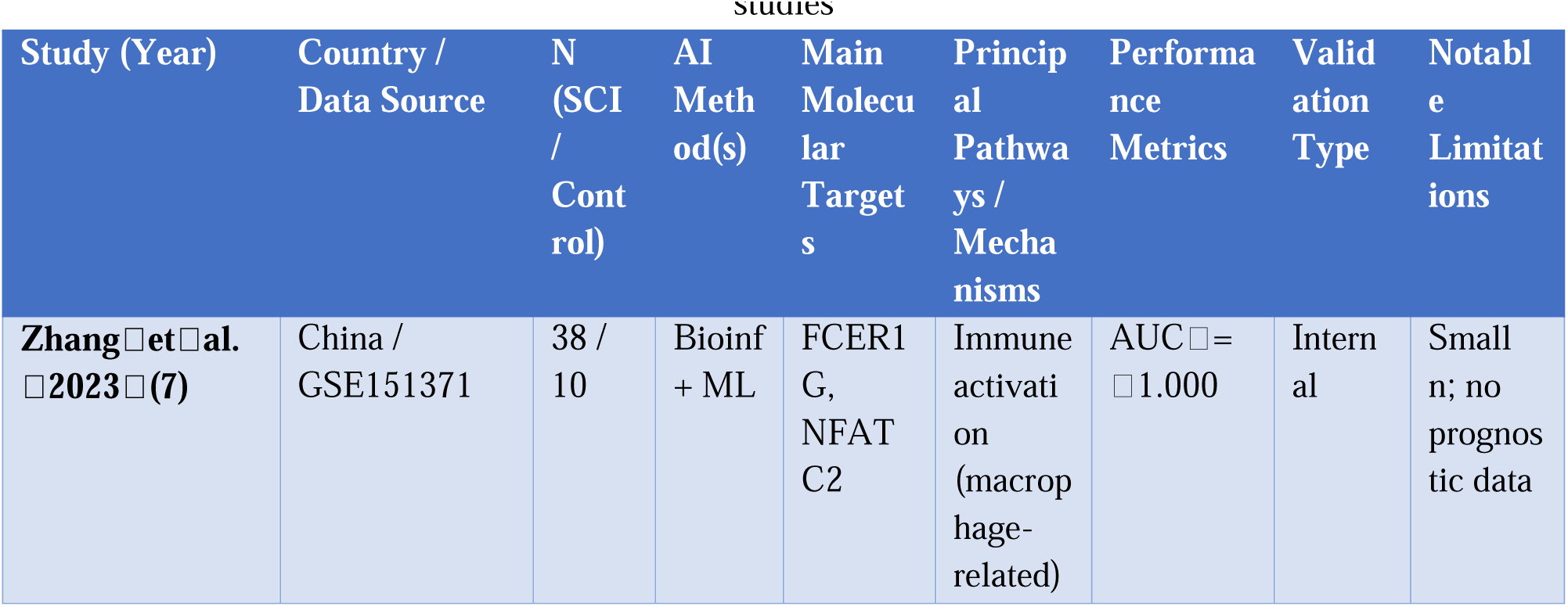
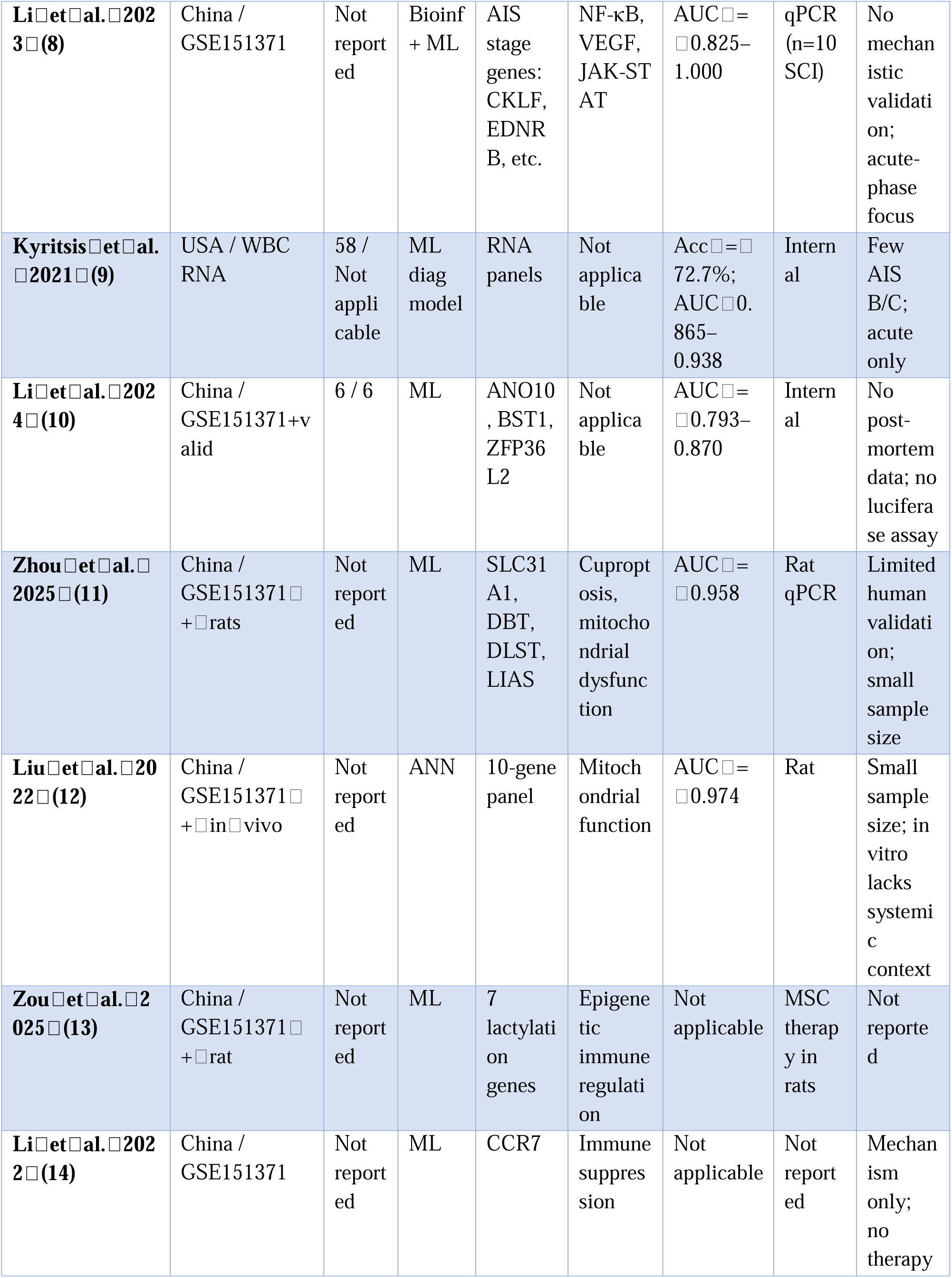
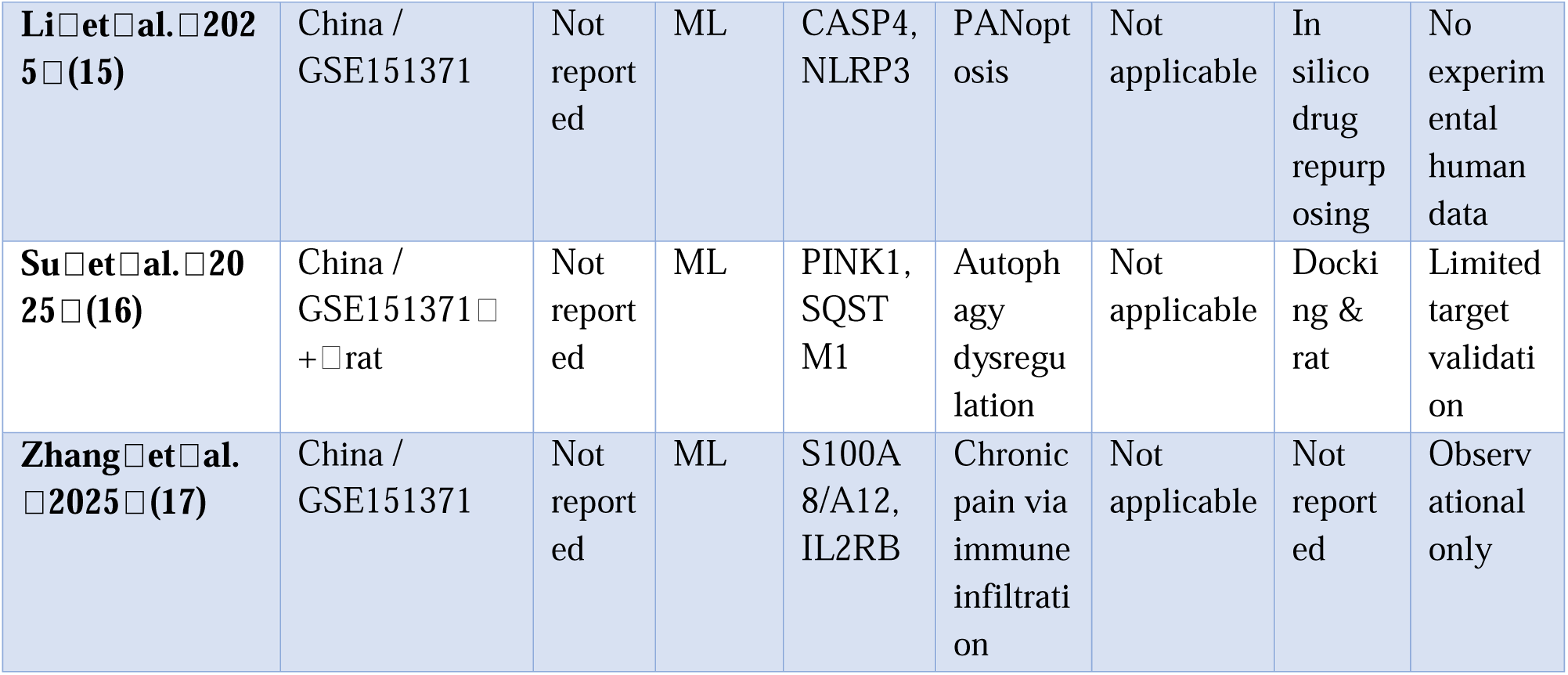
summary of data sources, AI models, pathways, and key metrics across included SCI–AI studies.

### Aggregate Summary of Data Sources, AI Models, and Pathways

Across the 11 included studies (7–17):

- Data type: 8/11 (72%) used blood-derived transcriptomic datasets, predominantly GSE151371; 3/11 (27%) used fresh RNA-seq from white blood cells, post-mortem cord tissue, or animal SCI models.
- AI algorithms: 7/11 (64%) used artificial neural networks (ANN) or ANN-based hybrid models; others employed support vector machines (SVM), random forests (RF), or ensemble methods.
- Pathway categories: 9/11 (82%) reported immune-related signaling (NF-κB, JAK-STAT, VEGF, toll-like receptor); 5/11 (45%) identified cell death programs (e.g., PANoptosis, cuproptosis, autophagy); 4/11 (36%) implicated mitochondrial metabolism.
- Validation: Only 3/11 studies used independent datasets beyond GSE151371; AUCs in these were more modest (0.79–0.87) compared to GSE151371-based models (often ≥□0.95).

This aggregated overview, absent in the original text, underscores dominant trends, recurrent biology, and algorithm choices.

### Diagnostic & Severity Classification Outcomes

Six studies primarily developed AI biomarker panels to diagnose SCI or classify injury severity (7–12). Most derived features from GSE151371 and applied machine learning for feature selection/classification. ***(Table 2.)***

- Performance: AUC values ranged from 0.793 to 1.000. Notably, *Zhang*□*et*□*al.*□(7) linked FCER1G and NFATC2 expression to macrophage-driven inflammation and achieved an AUC of 1.000 in their combined model.
- *Li*□*et*□*al.*□(8) created distinct gene sets for SCI diagnosis (e.g., CKLF, EDNRB, FCER1G) and AIS grades A/D, achieving AUCs of 0.825–1.000 in small qPCR cohorts (n□=□10 SCI).
- *Kyritsis*□*et*□*al.*□(9) used white blood cell RNA profiles to classify AIS grades with 72.7% accuracy (AUC 0.865–0.938), highlighting more moderate metrics in independent cohorts.
- *Li*□*et*□*al.*□(10) identified ANO10, BST1, ZFP36L2 with AUCs 0.793–0.870;
- *Zhou*□*et*□*al.*□(11) implicated cuproptosis gene SLC31A1 (AUC□=□0.958);
- *Liu*□*et*□*al.*□(12) used a 10-gene ANN yielding AUC□=□0.974 and validated with ZnO nanoparticles in rats.

**Table 2.**
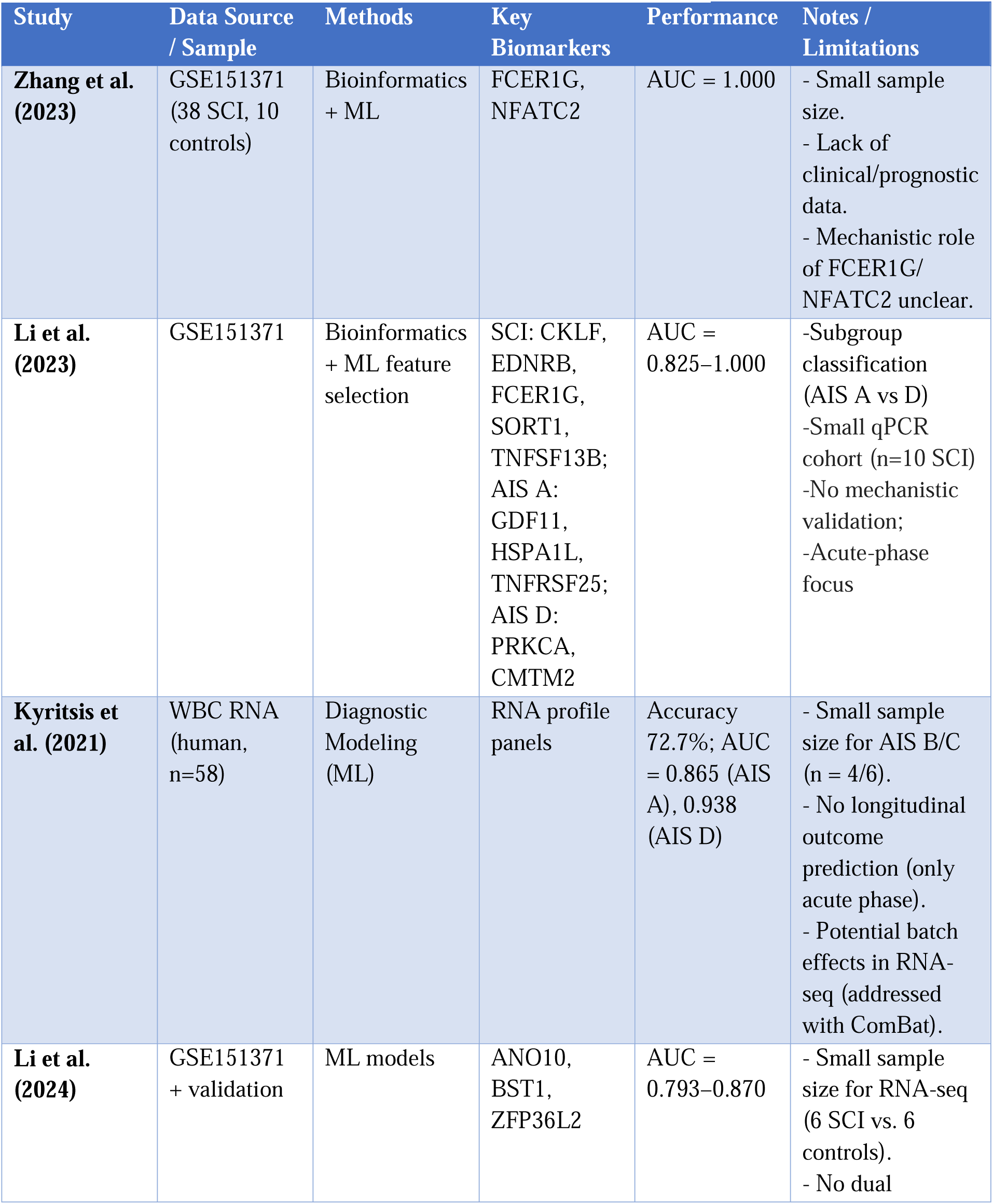
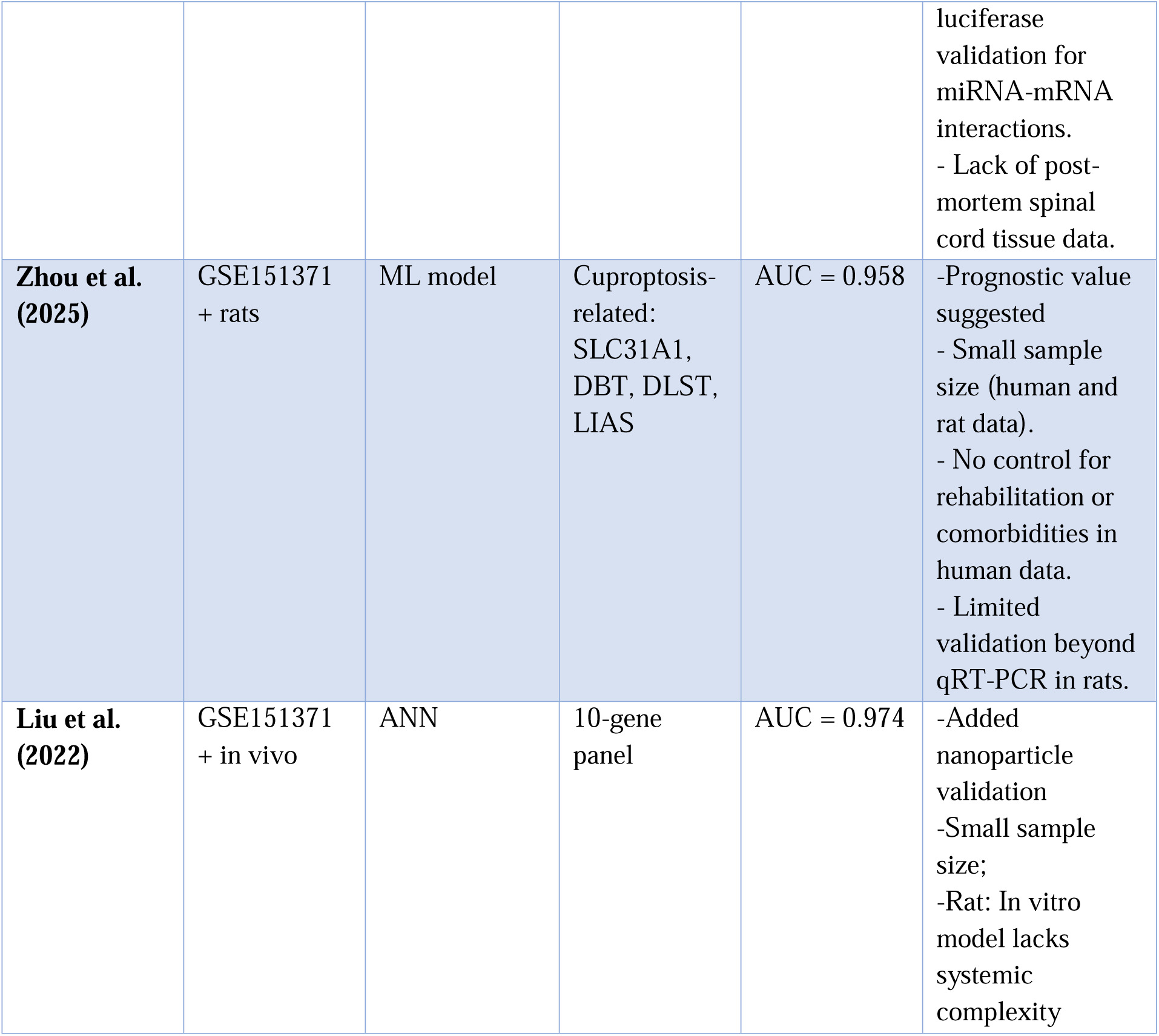
Diagnostic & Severity Classification Outcomes.

Bias consideration: Heavy GSE151371 use introduces cohort-specific batch effects; coupled with small n, this inflates AUCs and undermines generalizability.

### 2. Mechanistic & Pathophysiological Insights

Seven studies interrogated SCI biology via AI/bioinformatics (8,□11,□13–17):

- Immune dysregulation: *Li*□*et*□*al.*□(8) pinpointed NF-κB, VEGF, JAK-STAT; *Zou*□*et*□*al.*□(13) linked lactylation genes to immune infiltration; *Li*□*et*□*al.*□(14) tied CCR7 downregulation to immune deficiency.
- Cell death pathways: *Li*□*et*□*al.*□(15) highlighted PANoptosis (CASP4, NLRP3); *Zhou*□*et*□*al.*□(11) demonstrated mitochondrial injury via cuproptosis-related genes.
- Autophagy/metabolism: *Su*□*et*□*al.*□(16) reported PINK1/SQSTM1 imbalance;
- Chronic outcomes: *Zhang*□*et*□*al.*□(17) connected S100A8/A12 and IL2RB to neuropathic pain.

Recurrent themes included macrophage/T-cell infiltration, oxidative stress, mitochondrial dysfunction— implicated in both acute and chronic SCI phases.( Table 3)

**Table 3.**
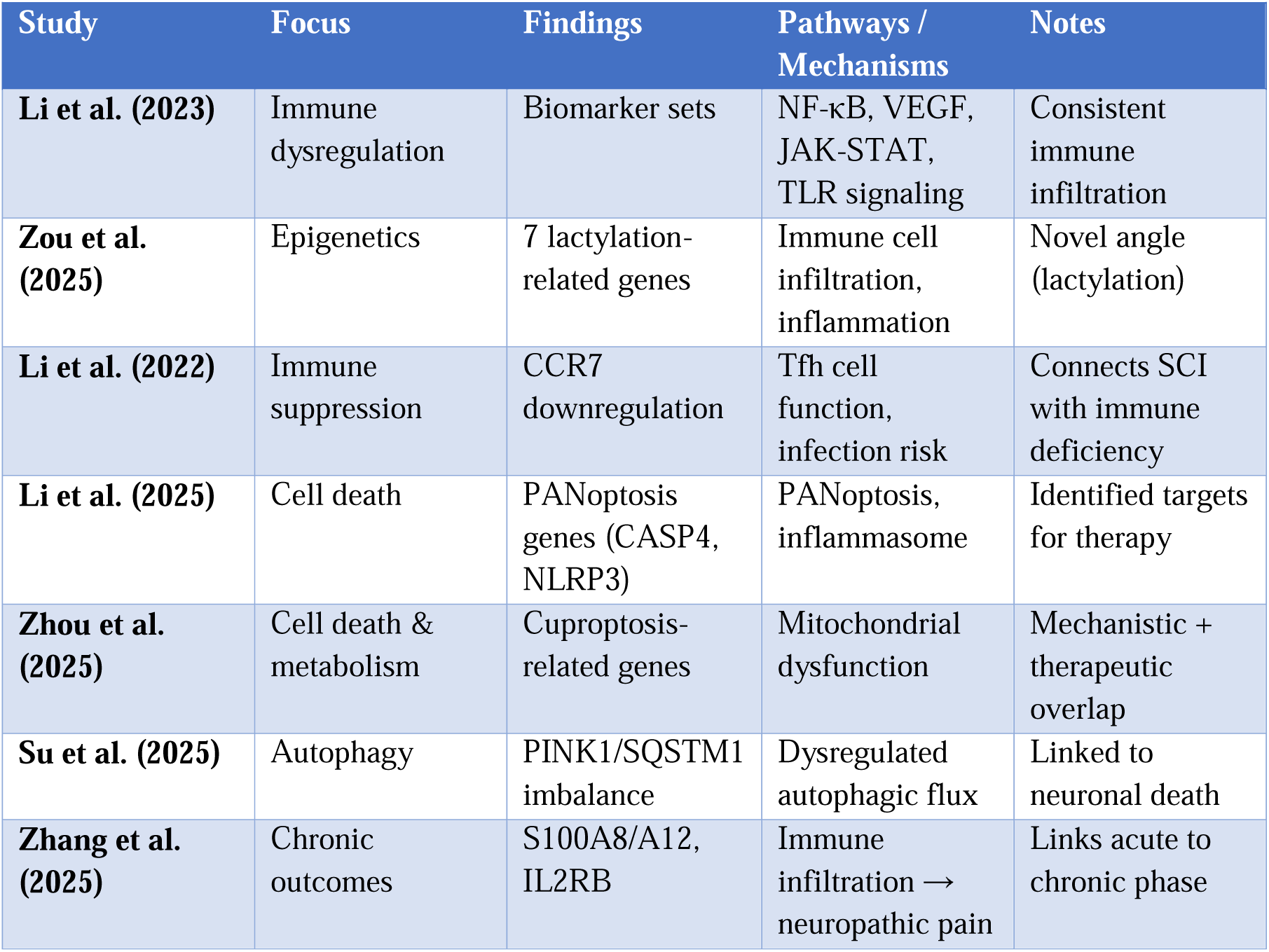
Mechanistic & Pathophysiological Insights.

### 3. Therapeutic Prediction & Experimental Validation

Five studies extended computational outputs to therapeutic hypotheses (11–13,□15–16)( Table 4):

- Drug repurposing: *Li*□*et*□*al.*□(15) → Emricasan, Alaproclate; *Su*□*et*□*al.*□(16) → Imatinib targeting PINK1 (ΔG□=□–10.9□kcal/mol).
- Nanoparticle/cellular therapies: *Liu*□*et*□*al.*□(12) → ZnO nanoparticles; *Zhou*□*et*□*al.*□(11) → mitochondrial protection; *Zou*□*et*□*al.*□(13) → mesenchymal stem cells.

**Table 4.**
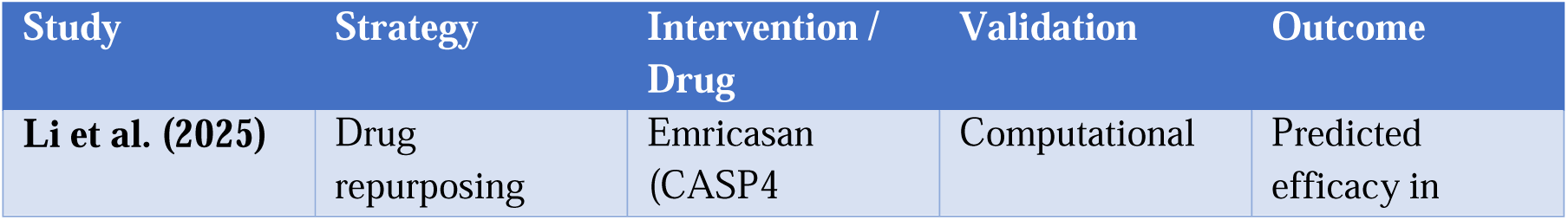
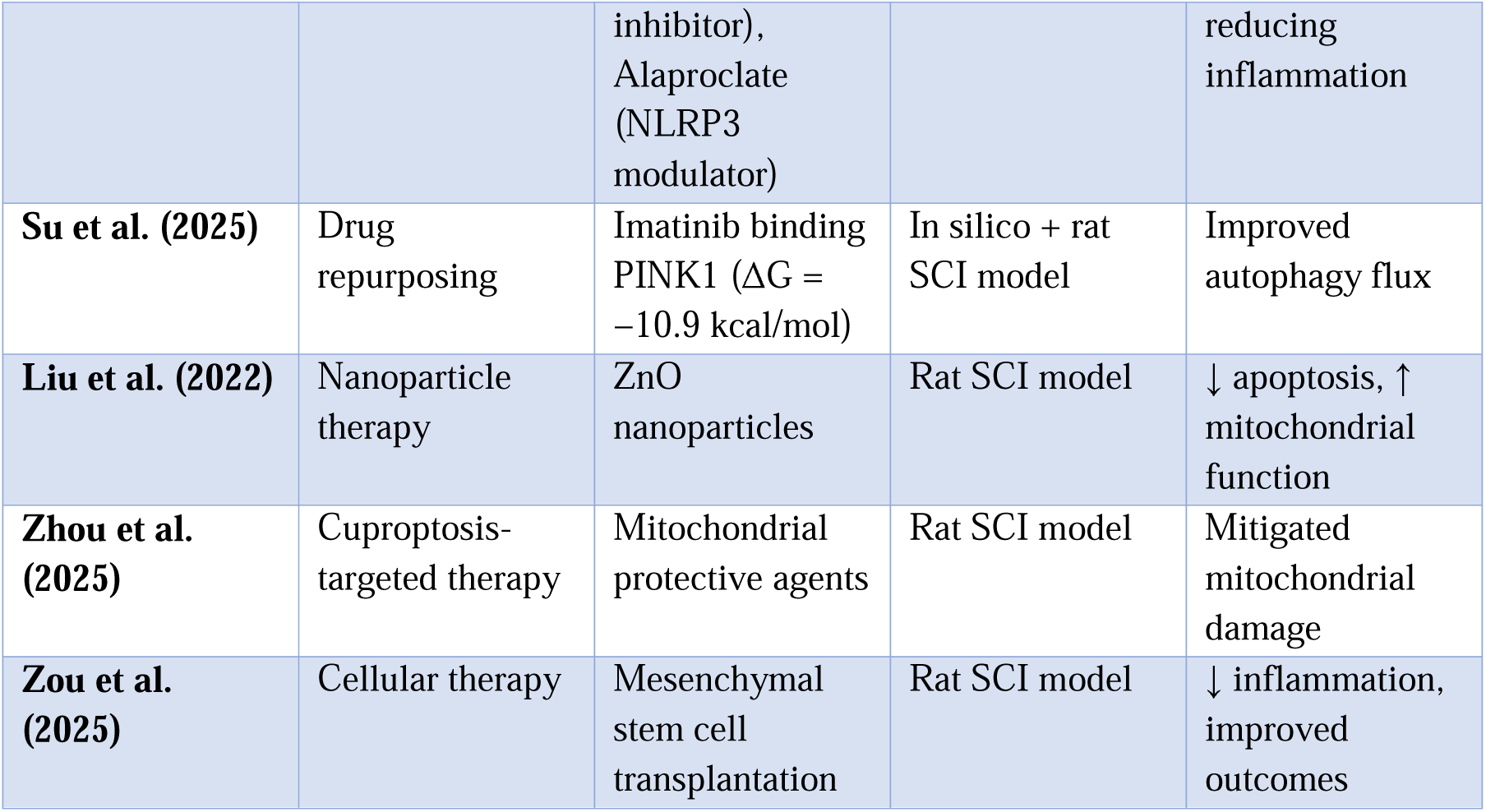
Therapeutic Prediction & Experimental Validation.

**Table 5.**
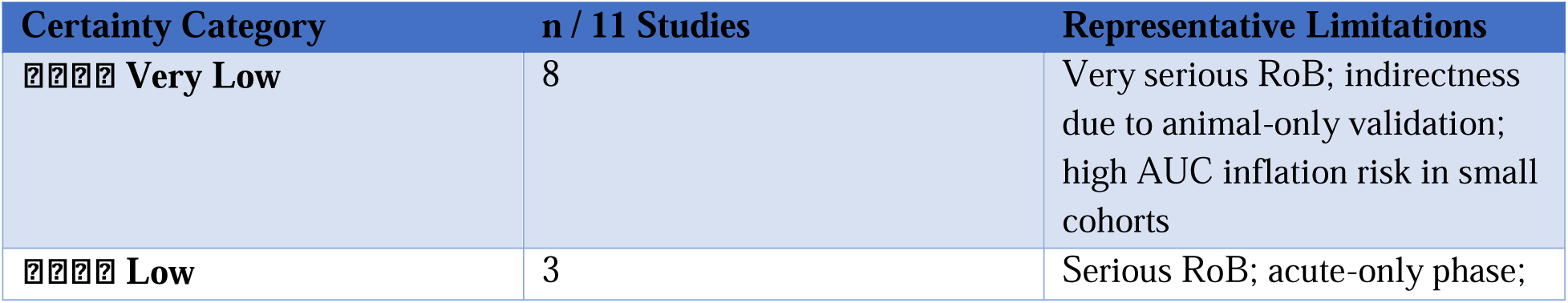
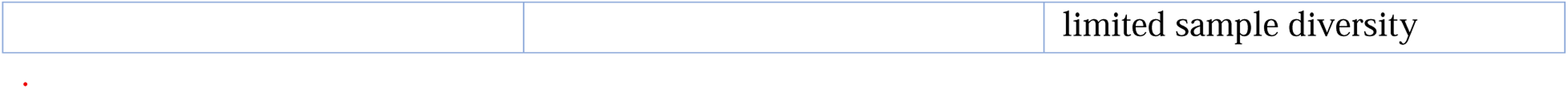
The GRADE Assessment.

Outcomes were positive in preclinical models (↓ inflammation/apoptosis, ↑ autophagy flux), but none have reached human trial phase.

### Impact of Dataset Dependence on Result Validity

Overrepresentation of GSE151371-derived models is a central limitation:

- Cohort homogeneity: Specific demographic and injury profile limits external validity.
- Single-lab processing: Risks laboratory-specific artefacts affecting gene expression signals.
- Metric inflation: Near-perfect AUCs not reproduced in heterogeneous datasets; independent test sets are rare.

These limitations are acknowledged across studies (7–12), but rarely quantified.

### Certainty of Evidence (GRADE) Assessment

Certainty of the evidence for all 11 included studies was systematically evaluated using the GRADE approach, integrating the risk-of-bias findings from our PROBAST assessments (Supplementary□4–5) with four additional domains: inconsistency, indirectness, imprecision, and publication bias. Downgrading was applied as *serious* (▼1) or *very serious* (▼2) according to predefined criteria ***(see supplementary 7***□***table***□***for full matrix)*.**

### Overall Certainty

Combining downgrades across domains, 8/11 studies were rated as Very Low Certainty (⍰⍰⍰⍰) and 3/11 studies as Low Certainty (⍰⍰⍰⍰). No included study achieved moderate or high certainty. The distribution and representative limitations are summarized in Table□5

### interpretation

The current body of AI-based SCI molecular pathway research remains predominantly hypothesis-generating. The pervasive use of a single transcriptomic dataset, reliance on bioinformatics without robust human validation, and absence of longitudinal clinical outcomes substantially limit confidence in the reported diagnostic, mechanistic, and therapeutic claims. These constraints underscore the urgent need for large, multi-center human cohorts and independent replication before clinical translation.

## Discussion

### 1. Certainty and Quality of Evidence (GRADE/PROBAST)

We applied the GRADE framework, supported by PROBAST risk-of-bias assessment ***(supplementary***□***7 tables),*** to all 11 included studies. Eight studies were rated very low certainty (⍰⍰⍰⍰) and three low certainties (⍰⍰⍰⍰). No study reached moderate or high certainty.

The most frequent downgrade reasons were:

- Serious risk of bias – Predominantly small, non-representative cohorts; heavy reliance on GSE151371; absence of independent multicenter validation.
- Indirectness – Therapeutic validation restricted to rodent models; acute-phase molecular snapshots without longitudinal human outcome linkage.
- Imprecision – Inflated AUC values without reporting confidence intervals; small event counts in model training and testing.
- Publication bias – Dataset reuse by overlapping author groups; lack of negative or neutral result reporting.

These limitations warrant interpreting current AI-driven molecular pathway mappings in SCI as hypothesis-generating rather than clinically definitive.

### 2. Summary of Main Findings

Diagnostic and Severity Classification

Six studies developed predictive models—mostly logistic regression, SVM, random forest, or ANN— using transcriptomic data (dominantly GSE151371) to detect SCI or grade its severity. Reported AUC values ranged from 0.79 to 1.00, with some single-gene models reporting near-perfect discrimination (e.g., FCER1G, NFATC2)□[1].

Li□et□al. stratified patients by AIS grade using separate biomarker panels (e.g., GDF11, HSPA1L, TNFRSF25 for AIS□A)□[2], and other recurrent markers included S100A8 and IL2RB□[3,4]. However, small validation cohorts (n□≤□10 in some qPCR sets) and repeated dataset use raise the risk of overfitting.

#### Mechanistic Insights

Seven studies shifted from classification to mechanistic mapping:

- Immune dysregulation :Macrophage polarization, T-cell suppression, neutrophil recruitment; NF-κB, VEGF, JAK-STAT, and Toll-like receptor signaling were repeatedly implicated□[2,5,8].
- Novel cell-death modalities :PANoptosis (CASP4, NLRP3) and cuproptosis (SLC31A1, DBT, DLST, LIAS) linked to mitochondrial impairment□[9,5].
- Metabolic and autophagy imbalance :PINK1/SQSTM1 disruption compromising autophagic flux□[10]; oxidative stress and mitochondrial metabolism as recurrent denominators.
- Chronic outcome links :S100A8/A12 and IL2RB implicated in immune infiltration–driven neuropathic pain□[11].

#### Therapeutic Predictions and Validation

Five studies used AI-derived targets to propose or test treatments:

- Drug repurposing :Emricasan (CASP4 inhibition), Alaproclate (NLRP3 modulation), Imatinib (PINK1 binding ΔG = –10.9□kcal/mol)□[9,10].
- Nanoparticle therapy :ZnO nanoparticles for mitochondrial protection□[6].
- Cell-based approaches :Mesenchymal stem-cell transplantation reducing inflammatory signatures□[7].

All validations remained preclinical (predominantly rodent SCI models), leaving an unaddressed human translational gap.

### 3. Comparison with Prior Literature

Our results align with prior AI-SCI reviews□[12,13], which reported promising diagnostic performance countered by limited cohort diversity. The recurrent identification of immune-metabolic intersections mirrors findings in traumatic brain injury and stroke, where AI-assisted transcriptomics have guided regenerative therapeutics□[14].

Outside SCI, integrating AI with CRISPR gene editing, single-cell omics, and bioengineered scaffolds has accelerated regenerative strategies. Conversely, Martín-Noguerol□et□al.□[15] highlight imaging-based AI in spinal pathology diagnosis, underscoring potential synergies between omics and imaging models yet to be exploited in SCI.

### 4. Limitations of Included Studies

Key constraints identified across the evidence base include:

- Dataset dependence :Overreliance on GSE151371 reduces external validity.
- Small sample size :Particularly acute in AIS□B/C subgroups; diminishes statistical power.
- Restricted temporal scope :Acute-phase molecular states without chronic follow-up correlations.
- Preclinical validation bias :Therapeutic efficacy assessed almost exclusively in rodents.
- Selective reporting :Pathways with modest statistical signals likely under-reporte

### 5. Implications for Regenerative Medicine

The overlap between AI-prioritized biomarkers and regenerative pathway targets provides tangible translational opportunities:

- Targeting immune–metabolic crosstalk :PANoptosis and cuproptosis nodes as junctions where inflammation and metabolism intersect.
- Enhancing autophagy and mitochondrial protection :Potential to improve neuronal survival and axonal regrowth.
- Combination strategies :Embedding AI-predicted molecular targets into biomaterial scaffolds, stem-cell constructs, or exosome-mediated delivery systems.

For regenerative translation, early-phase human trials should explicitly link molecular biomarker modulation to both functional recovery (e.g., motor scores) and neuroimaging changes (e.g., DTI metrics).

### 6. Future Directions

To bridge the current discovery-to-bedside gap, we recommend:

1. Building multi-center, multi-omics cohorts :Standardized sampling across acute, subacute, and chronic phases; integrating transcriptomics, proteomics, metabolomics, and advanced imaging.
2. Expanding diversity :Recruiting broader age ranges, injury severities, and geographic backgrounds to improve generalizability.
3. Longitudinal correlation :Directly linking early molecular signatures with chronic functional and clinical outcomes.
4. Advancing translational research :Moving AI-predicted drugs and regenerative constructs into adaptive early-phase clinical trials.
5. Ensuring transparent reporting :Publishing replication attempts, null results, and dataset metadata to mitigate publication and analytic biases.

## Conclusion

This systematic review shows that artificial intelligence, particularly machine learning–driven bioinformatics, can reliably detect transcriptomic signatures linked to spinal cord injury diagnosis, severity grading, and mechanistic profiling. Across the 11 included studies, recurrent biomarkers such as FCER1G, S100A8, IL2RB, PINK1, and genes involved in novel cell-death programs (PANoptosis, cuproptosis) were consistently prioritized, pointing to immune infiltration, oxidative stress, and mitochondrial metabolism as central to SCI pathogenesis. Therapeutic predictions derived from these models—including repurposed drugs (Emricasan, Alaproclate, Imatinib), nanoparticle-based mitochondrial protection, and mesenchymal stem-cell transplantation—produced encouraging preclinical results but remain untested in human cohorts. Despite high reported diagnostic accuracies (AUC□=□0.79–1.00), methodological weaknesses such as over-reliance on a single dataset (GSE151371), small and homogeneous validation cohorts, acute-phase bias, and absence of longitudinal clinical correlation substantially limit generalizability. To bridge the translational gap, future research must focus on multi-center, multi-omics, and longitudinal designs, with rigorous external validation and integration of imaging and functional outcomes. Advancing the most promising AI-predicted targets into early-phase clinical trials is critical. Real-world application will depend on moving beyond computational discovery to clinically verified interventions that improve neurological recovery and quality of life for SCI patients.

## Data Availability

All data produced in the present work are contained in the manuscript

