## Supplementory for "Artificial Intelligence Models for Predicting Molecular Pathway Activity in Spinal Cord Injury: A Systematic Review"

Conceptualization: MM1; Data curation: MM1, MJ, BR,RT; Investigation: MJ, BR,IM; Methodology: MM1; Supervision: FF,SO2,AZ; Visualization: MM1,MM2; Writing – original draft: MM1, MJ, BR,SO1,RT,IM,MM2 Writing – review and editing: FF,SO2,AZ. All authors reviewed and confirmed the final draft of this manuscript for submission.
