## Supplementory for "Artificial Intelligence Models for Predicting Molecular Pathway Activity in Spinal Cord Injury: A Systematic Review"

Supplementary Methods – Certainty Assessment

Certainty of evidence was rated per GRADE domains: *(i)* Risk of bias via PROBAST findings, *(ii)* Inconsistency across comparable studies, *(iii)* Indirectness of population/intervention/outcome vs. review question, *(iv)* Imprecision (sample size, CI, overfitting risk), *(v)* Publication bias.

Downgrading: Serious = ▼1; Very serious = ▼2. Most studies were downgraded in ≥3 domains due to small sample sizes, heavy dependence on GSE151371, and absence of large independent validations. Overall, 8/11 studies were rated Very Low Certainty, and 3/11 as Low Certainty.

**Risk of Bias:**

Eight of eleven studies were downgraded by two levels for very serious risk of bias, primarily due to small sample sizes, overreliance on the same blood‑derived transcriptomic dataset (GSE151371), and absence of independent external validation. The remaining three studies were downgraded by one level for serious limitations such as restricted sub‑cohorts (e.g., AIS B/C n<10) or lack of mechanistic confirmation beyond bioinformatics predictions.

**Inconsistency:**

Although immune‑related pathways (e.g., NF‑κB, JAK‑STAT) and recurrent biomarkers (e.g., FCER1G, IL2RB, S100A8) were identified in multiple datasets, heterogeneity existed in the magnitude and specificity of reported effects. Five studies were downgraded for inconsistency, reflecting variability in gene panels and model performance not fully explained by population or methodological differences.

**Indirectness:**

Seven studies were downgraded (one to two levels) for using predominantly animal models or highly specific acute‑phase cohorts without direct linkage to long‑term functional outcomes. Particularly, therapeutic prediction studies with only rodent validation (e.g., ZnO nanoparticles, mitochondrial protective agents) were rated as having high indirectness to clinical applicability.

**Imprecision:**

Ten studies were downgraded for imprecision, largely due to small effective sample sizes, absence of confidence intervals, and possible overfitting indicated by near‑perfect AUC values (0.97–1.00) in minimal validation cohorts.

**Publication Bias:**

Potential for selective reporting was inferred in six studies from dataset reuse within overlapping research groups and absence of negative outcome reporting. This domain was downgraded for these studies.

**Overall Certainty:**

Combining downgrades across domains, **8/11 studies** were rated as **Very Low Certainty (✦✧✧✧)** and **3/11 studies** as **Low Certainty (✦✦✧✧)**. No included study achieved moderate or high certainty.

َ

| **Study (Year)** | **RoB (PROBAST)** | **Inconsistency** | **Indirectness** | **Imprecision** | **Publication Bias** | **Overall Certainty** |
| --- | --- | --- | --- | --- | --- | --- |
| Zhang 2023 | ▼2 – small sample (38+10), single dataset, no clinical outcomes | ▼1 – overlap in immune genes but diff. emphasis | ▼1 – acute phase only | ▼1 – no CI, possible AUC inflation | ▼1 – dataset reuse in field | ✦✧✧✧ Very Low |
| Li 2023 | ▼2 – small qPCR (n=10), no mech. validation | ▼0 – consistent immune-related findings | ▼1 – acute only | ▼1 – CI absent; AUC 1.000 risk overfit | ▼1 | ✦✧✧✧ Very Low |
| Kyritsis 2021 | ▼1 – small AIS B/C subgroup, BEs | ▼1 – moderate variation vs other studies | ▼0 – direct human | ▼1 – low subgroup N | 0 – no evident pub bias | ✦✦✧✧ Low |
| Li 2024 | ▼2 – n=6 vs n=6, no tissue confirm | ▼1 – markers differ partly | ▼1 – confined demographics | ▼1 – CI absent | ▼1 – same dataset | ✦✧✧✧ Very Low |
| **Zhou 2025** (Diag/Prog) | ▼2 – small mixed human + rat | ▼0 – aligns with other cuproptosis data | ▼1 – partly animal | ▼1 – low N | ▼1 | ✦✧✧✧ Very Low |
| **Liu 2022** (Diag/Ther) | ▼2 – unclear small human, heavy rat | ▼0 – overlaps | ▼2 – animal predominant | ▼1 | ▼1 | ✦✧✧✧ Very Low |
| Zou 2025 | ▼1 – data source unclear, likely small human | ▼1 – novelty (lactylation) but overlaps pathways | ▼1 – uncertain human applicability | ▼1 | ▼1 | ✦✧✧✧ Very Low |
| Li 2022 | ▼1 – limited CCR7 data, small N | ▼0 – immune suppression unique | ▼1 – indirect to clinical outcome | ▼1 | 0 | ✦✦✧✧ Low |
| **Li 2025** (PANoptosis) | ▼1 – bioinfo only, no in vivo human | ▼0 – novel mechanism but fits data | ▼1 – indirect to humans | ▼1 | ▼1 | ✦✧✧✧ Very Low |
| Su 2025 | ▼1 – modeled + rat | ▼0 – consistent autophagy dysreg. | ▼2 – animal heavy validation | ▼1 | ▼1 | ✦✧✧✧ Very Low |
| Zhang 2025 | ▼1 – bioinfo chronic pain, no large N | ▼1 – overlaps immune pathways | ▼1 – indirect to acute SCI diag/severity | ▼1 | 0 | ✦✦✧✧ Low |
