## Supplementory for "Artificial Intelligence Models for Predicting Molecular Pathway Activity in Spinal Cord Injury: A Systematic Review"

| Item | Checklist item | Location where item is reported |
| --- | --- | --- |
| 1 | Identify the report as a systematic review. | Title page identified as systematic review |
| 2 | See the PRISMA 2020 for Abstracts checklist. | Structured abstract (Background, Objectives, Methods, Results, Conclusions) |
| 3 | Describe the rationale for the review. | Introduction (Background, comparison with prior reviews) |
| 4 | Provide an explicit statement of the objectives. | Abstract + Aim of the Present Review |
| 5 | Specify inclusion and exclusion criteria. | Eligibility Criteria section |
| 6 | Specify all databases and sources searched. | Search Strategy section (PubMed, Scopus, Web of Science, Google Scholar, reference lists) |
| 7 | Present the full search strategies. | Search Strategy + Supplementary 2 (full syntax) |
| 8 | Specify methods used for study selection. | Screening Process (two reviewers independently, consensus resolution) |
| 9 | Specify methods used for data collection. | Data Extraction section |
| 10a | List and define all outcomes for which data were sought. | Outcomes section (primary: AI model performance; secondary: pathways, biomarkers, translational recs) |
| 10b | List and define all other variables sought. | Data Extraction section (bibliographic details, AI methods, datasets, assumptions) |
| 11 | Specify methods used to assess risk of bias. | Risk of Bias Assessment (PROBAST tool) |
| 12 | Specify effect measures used. | Outcomes section (AUC, accuracy, precision, recall, F1, specificity, sensitivity, effect size) |
| 13a | Describe processes to decide synthesis eligibility. | Methods — grouped by diagnostic, mechanistic, therapeutic outcomes |
| 13b | Describe methods for data preparation. | Not explicitly described (data prep methods not detailed) |
| 13c | Describe methods for tabulating/displaying results. | Tables 1–4 + Figure 2 (PRISMA flowchart) |
| 13d | Describe methods used to synthesize results. | Narrative synthesis (meta-analysis not performed due to heterogeneity) |
| 13e | Describe methods to explore heterogeneity. | Not performed (heterogeneity acknowledged) |
| 13f | Describe sensitivity analyses conducted. | Not applicable (no meta-analysis) |
| 14 | Describe methods to assess reporting bias. | Certainty of Evidence (GRADE) + Discussion (publication bias) |
| 15 | Describe methods to assess certainty of evidence. | Certainty of Evidence (GRADE) Assessment |
| 16a | Describe results of search and selection process. | Results + Figure 2 (PRISMA flowchart) |
| 16b | Cite excluded studies with reasons. | Supplementary 3a & 4a (exclusion reasons) |
| 17 | Cite each included study and present its characteristics. | Results + Table 1 (study characteristics) |
| 18 | Present assessments of risk of bias. | Figure 1 (PROBAST risk of bias) + text |
| 19 | Present results of individual studies. | Tables 1–3 (study outcomes, effect estimates) |
| 20a | Summarise characteristics and RoB among contributing studies. | Results (aggregate summaries, diagnostic outcomes, mechanistic insights, therapeutic predictions) |
| 20b | Present results of statistical syntheses. | Not applicable (no meta-analysis) |
| 20c | Present results of investigations of heterogeneity. | Discussed (dataset dependence, overfitting, etc.) |
| 20d | Present results of sensitivity analyses. | Not applicable (no sensitivity analyses) |
| 21 | Present risk of bias due to missing results. | Certainty assessment + Discussion (publication bias) |
| 22 | Present certainty of evidence assessments. | GRADE table + Results + Discussion |
| 23a | Provide a general interpretation of results. | Discussion (summary of findings, comparison with prior reviews) |
| 23b | Discuss limitations of the evidence. | Discussion (limitations of included studies) |
| 23c | Discuss limitations of the review process. | Discussion (limitations of review processes) |
| 23d | Discuss implications for practice, policy, research. | Discussion (implications for regenerative medicine, policy, research) |
| 24a | Provide registration information. | Methods (PROSPERO registration CRD420251115723) |
| 24b | Indicate where the protocol can be accessed. | Registered in PROSPERO; no external link provided |
| 24c | Describe and explain protocol amendments. | Amendments: meta-analysis not performed due to heterogeneity |
| 25 | Describe sources of support. | Before references |
| 26 | Declare competing interests. | Before references |
| 27 | Report availability of data, code, materials. | Supplementary files (search strategy, exclusion sheets, extracted data, GRADE tables) |
