## Supplementory for "Artificial Intelligence Models for Predicting Molecular Pathway Activity in Spinal Cord Injury: A Systematic Review"

Search Strategy:

**SCOPUS (n=45):**

  TITLE-ABS-KEY ("spinal cord injury" OR "SCI" OR "spinal trauma" OR "myelopathy")

  AND

  TITLE-ABS-KEY ("artificial intelligence" OR "AI" OR "machine learning" OR "deep learning" OR "neural network" OR "predictive model" OR "random forest" OR "support vector machine" OR "SVM")

  AND

  TITLE-ABS-KEY ("molecular pathway" OR "signaling pathway" OR "gene network" OR "transcriptomic" OR "proteomic" OR "metabolomic" OR "bioinformatic" OR "systems biology")

**WOS (n=22):**

TS= ("spinal cord injury" OR "SCI" OR "spinal trauma")

AND

TS= ("artificial intelligence" OR "machine learning" OR "deep learning" OR "neural network")

AND

TS= ("molecular pathway" OR "signaling cascade" OR "transcriptome" OR "proteome" OR "metabolome" OR "bioinformatics")

**PUBMED (n=19):**

  ("spinal cord injury"[MeSH] OR "spinal cord injuries"[Title/Abstract] OR "SCI"[Title/Abstract])

  AND

  ("artificial intelligence"[MeSH] OR "machine learning"[Title/Abstract] OR "deep learning"[Title/Abstract] OR "neural networks"[Title/Abstract])

  AND

  ("signal transduction"[MeSH] OR "molecular pathways"[Title/Abstract] OR "gene regulatory networks"[Title/Abstract] OR "transcriptomics"[Title/Abstract])
