## Supplementory for "Artificial Intelligence Models for Predicting Molecular Pathway Activity in Spinal Cord Injury: A Systematic Review"

PRISMA Checklist:

1. Title: Predictive Artificial Intelligence Modeling of Molecular Signaling Pathways in Spinal Cord Injury

2. Abstract:

3. Introduction (Rationale): Spinal cord injury (SCI) is a severe condition affecting millions worldwide, leading to profound motor and sensory impairments. Most SCI cases are caused by traumatic events such as accidents and falls. The injury process is complex, involving immediate mechanical damage and ongoing harm from inflammation, oxidative stress, and scar formation, which hinder recovery. Despite progress in AI and molecular signaling research, a gap exists in synthesizing these findings into a cohesive framework to guide SCI treatment strategies. This systematic review aims to consolidate existing research, assess the accuracy of AI models, and identify key molecular pathways for targeted therapies. By evaluating AI modeling in SCI, the review will enhance our understanding of how AI can improve therapeutic approaches and outcomes.

4 . Introduction (Objectives): Molecular signaling pathways are central to the SCI pathophysiology, governing cellular responses, such as inflammation, cell survival, and neuronal regeneration. Dysregulated pathways, including the activation of pro-inflammatory cytokines and the involvement of glial cells, exacerbate tissue damage, ultimately leading to neuronal death and axonal degeneration. Artificial Intelligence (AI) has emerged as a transformative tool in SCI research, enabling the analysis of large datasets to derive predictive models. AI helps identify biomarkers and predict the activation of molecular pathways post-injury, offering insights into recovery or further damage.

5. Methods (Eligibility Criteria): Inclusion criteria comprised peer-reviewed articles that applied AI techniques to predict and model molecular pathway activity in spinal cord injury (SCI). The study designs contain a cross-sectional design with an in vivo/ in vitro follow-up, retrospective analysis of a prospective cohort, registered clinical trial, and biomarker validation through a prospective cohort, bioinformatics analysis of a retrospective cohort and molecular signaling analysis, cross-sectional observational study, retrospective transcriptome analysis, and qPCR validation cross-sectional observational study, bioinformatics analysis and experimental validation, experimental validation, animal model follow up, and cross sectional research using animal models. The minimum follow-up was 48 hours after injury.

6. Information Sources: We operated various databases from Google Scholar, PubMed, and Web of Science. The datasets from the references contain GEO datasets, GSE 151371, GSE 151317 SCI blood, Track SCI study, and GSE 226238.

7. Search Strategy: spinal cord injury or SCI or spinal trauma or myelopathy and artificial intelligence or AI or machine learning or deep learning or neural network or predictive model or random forest or support vector machine or SVM and molecular pathway or signaling pathway or gene network or transcriptomic or proteomic or metabolomic or bioinformatic or systems biology

8. Selection Process: We have utilized Rayyan for inclusion and exclusion criteria, three reviewers screened each record and each report independently.

9. Data Collection Process: Overview and PROSPERO, review data, screening, full text screening, data extraction

10. Data Items A.(Outcomes): Autophagy-related biomarkers and potential therapeutic drugs for spinal cord injury, offering promising tools for early diagnosis and targeted treatment in clinical settings, novel biomarkers for early SCI diagnosis, highlighting immune response modulation for therapeutic targeting, potential of biomarkers for SCI diagno, sis and therapeutic targets, potential for identifying new biomarkers and therapeutic targets for chronic pain post-SCI, High diagnostic potential for SCI-IDS using peripheral blood markers, high potential for early diagnosis and grading of SCI using blood biomarkers, Blood RNA profiles could potentially be used in clinical settings to predict SCI severity and inform treatment decisions, the identification of PANoptosis-related genes provides new insights into the molecular mechanisms of SCI. Targeting these genes with small molecule inhibitors may offer a potential therapeutic strategy for spinal cord injury, suggest a new therapeutic avenue for SCI treatment via ZnO NPs, and potentially target lactate metabolism in SCI therapy, offering personalized treatment strategies.

10.Data items B.(Other Variables): biomarkers including " NLRC4, PINK1, VAMP3, FCER1G, NFATC2, ANO10, BST1, ZFP36L2, IL2RB, S100A8, NKG7, S100A12, CCR7, SCI General:CKLF, EDNRB, FCER1G, SORT1, TNFSF13B, AIS A Grade: GDF11, HSPA1L, AIS D Grade:PRKCA, CMTM2, Metalloproteinase-8 (MMP8), Haptoglobin (HP), Kininogen-1 (KNG1), MCEMP1, SOD1, SOD2, Bax, Bcl2, IL-4, IL-10, IL-1β, TNF-α , SLC31A1, DBT, DLST, LIAS, LSP1, XRCC4, HSDL2, HNRNPH1, RPL14, IKZF1, TP53", signaling pathways "Autophagy, apoptosis, neurodegeneration, NOD-like receptor signaling, Immune system processes, Th1/Th2 cell differentiation, T-cell receptor signaling, T cell receptor signaling, NF-κB signaling, immune-inflammatory pathways, T cell activation, immune response pathways, neuroinflammation, T-cell receptor signaling, chemokine pathways, Immune suppression, CCR7 Cytokine-cytokine receptor interaction,VEGF, JAK-STAT, Toll-like receptor, sphingolipid signaling Immune response, cellular secretion and localization, Inflammatory signaling, pyroptosis, apoptosis, necroptosis, PI3K/Akt signaling pathway (both human and animal data), cuproptosis-related (TCA cycle, mitochondrial metabolism) and Ribosome, HIF-1 signaling pathway, T cell receptor signaling, NF-kB signaling, Th1/Th2 cell differentiation, mTOR signaling, ", artificial intelligence models " Machine learning algorithms (LASSO, Random Forest), Machine learning models, including Random Forest and LASSO for feature selection + PPI, SVM, LASSO regression, SVM-RFE, Random Forest, R (version 4.2.0), Cytoscape, STRING, clusterProfiler, ConsensusClusterPlus, CIBERSORT, GSVA,R packages (limma, clusterProfiler, CIBERSORT, ggplot2, WGCNA),R (edgeR, limma, CIBERSORTx), Illumina RNA-seq, DESeq2 (RNA-seq analysis), R packages (pheatmap, EnhancedVolcano), "limma" package, "pheatmap", "reshape2", "ggpubr", "GSVA", "GSEABase", "ConsensusClusterPlus", "Venn", "pROC", "GraphPad Prism8", R packages: "limma" for gene expression analysis, CIBERSORT for immune cell infiltration, DESeq2 for RNA-seq analysis "

11. Study Risk of Bias Assessment: overall judgement of risk of bias: high, overall judgement of applicability concerns: low

12. Effect Measures: We gathered data on report: author, year, and source of publication, the study: sample characteristics, social demography, and definition and criteria used for spinal cord injury, the participants: social situation, time-lapsed since SCI, history of SCI, current neurological status, current treatments for SCI, the research design and features: sampling mechanism, treatment assignment mechanism, adherence, non-response, and length of follow-up, the intervention: type, duration, dose, timing, and mode of delivery

13. A. Synthesis Methods (Eligibility for Synthesis): In cases where the means, number of participants, and test-statistics were reported and there was the opportunity to include results in a meta-analysis, we calculated the standard deviations, assuming the standard deviation for each of the two groups (intervention and control).

13.B.Synthesis Methods (Preparing for Synthesis): Given the complexity of the interventions being investigated, we attempted to categorize the included interventions along four dimensions. (1)Operate on spinal cord injury models (2)Consume human data or clinical datasets. (3)Apply molecular mechanisms and/or stem cell interventions in SCI models. (4)Utilize predictive applications of artificial intelligence in prognostic therapeutic procedures in human SCI using molecular mechanisms.

(1)Spinal Cord Injury models

(2) human data or clinical datasets

(3)molecular mechanisms and/or stem cell interventions in SCI models

(4)The predictive applications of artificial intelligence

13.C. Synthesis Methods (Tabulation and Graphical Methods):

| Article | Number of Cases | Injury Location | Datasets | AI Model | Main Signaling Pathways | Main Findings |
|---|---|---|---|---|---|---|
| *Identification of Autophagy-Related Genes in Patients with Acute Spinal Cord Injury and Analysis of Potential Therapeutic Targets (Xiaochen Su et a,- 2024)* | 10 healthy controls, 38 SCI patients | Thoracic (T9-T10 in rats) | GEO dataset (GSE151371) | Machine learning algorithms (LASSO, Random Forest) | Autophagy, apoptosis, neurodegeneration, NOD-like receptor signaling | Pathways related to autophagy, apoptosis, neurodegeneration, and NOD-like receptor signaling were significantly enriched. |
| *Comprehensive landscape of immune-based cclassifiers related to early diagnosis and macrophage M1 in spinal cord injury (Zhang et al., 2023*) | 10 healthy individuals 38 the SCI group + 10 SCI, 8 HC in external validation | Cervical, Thoracic, Lumbar in GSE151371 T3–T12 in human cohort T10 in rats | GEO database (GSE151371), Xi-Jing Hospital cohort | Machine learning models, including Random Forest and LASSO for feature selection + PPI | Immune system processes, Th1/Th2 cell differentiation, and T-cell receptor signaling | identified immune-related genes (FCER1G and NFATC2) as biomarkers for early diagnosis of SCI. Immune dysregulation, including an increase in myeloid and a decrease in lymphoid post-SCI. |
| *Screening biomarkers for spinal cord injury using weighted gene co-expression network analysis* | 16 SCI patients 16 healthy controls | AIS grades A–D for GSE151371 dataset | GSE151371 dataset (public GEO database) | Machine learning algorithms (LASSO and SVM-RFE) | T cell receptor signaling, NF-κB signaling, immune-inflammatory pathways | Immune-inflammatory pathways are significantly altered in acute SCI - immune cell composition changes correlate with biomarkers |

| | | | | | | |
|---|---|---|---|---|---|---|
| *and machine learning (Li et al., 2024)* | | | | | | |
| Integrated bioinformatics analysis of the effects of chronic pain on patients with spinal cord injury (Zhang et al., 2025) | GSE151371: 38 SCI patients + 10 healthy controls GSE177034: 49 patients with chronic pain (post-SCI) | Spinal Cord "It didn't specify particularly." | GEO datasets GSE151371 (SCI blood) GSE177034 (chronic pain post-SCI) | LASSO regression SVM-RFE Random Forest | T cell activation, immune response pathways, and neuroinflammation | CCR7 is a strong diagnostic biomarker for acute SCI -Immune suppression mediated via Tfh cells associated with CCR7 downregulation post-SCI |
| *CCR7-mediated T follicular helper cell differentiation is associated with the pathogenesis and immune microenvironment of spinal cord injury-induced immune deficiency syndrome (Li et al., 2022)* | 58 (38 SCI, 10 healthy controls, 10 trauma controls) | AIS grades A–D in GSE151371 dataset check* | GEO dataset (GSE151371) | Logistic Regression (LR), LASSO, Random Forest Gradient Boosting Decision Tree (GBDT) XGBoost | - T-cell receptor signaling - Chemokine pathways -Immune suppression, CCR7 | Identified novel immune blood biomarkers for SCI diagnosis and AIS grade classification with high diagnostic accuracy Immune cell, including neutrophils and macrophages, increased in SCI patients vs HC. |
| *Identification of immunodiagnostic blood biomarkers associated with spinal cord injury* | 38 SCI patients + 10 trauma controls, 10 healthy controls External (qPCR) validation | Stratified by ASIA scale (Grades A–E) | GEO dataset GSE151371 | Multinomial logistic regression with LASSO regularization, Receiver Operating Characteristic (ROC) analysis | Cytokine-cytokine receptor interaction VEGF JAK-STAT Toll-like receptor Sphingolipid signaling | Blood RNA expression profiles from WBCs can predict SCI severity (AIS A and AIS D), providing a promising diagnostic biomarker |

| | | | | | | |
|---|---|---|---|---|---|---|
| *severity (Li et al., 2023)* | 1of 0 SCI patients, 3 trauma controls, 3 healthy controls | | | | | |
| Diagnostic blood RNA profiles for human acute spinal cord injury (Kyritsis et al., 2021) | 38 SCI patients, 10 healthy controls, 10 trauma controls | AIS grades A–D in the GSE151371 dataset | TRACK-SCI study (hospital registry)+ GEO database (GSE151371) | LASSO, SVM-RFE, XGBoost | Immune response, cellular secretion, and localization | Identification of 5 hub genes (CASP4, GSDMB, NAIP, NLRC4, NLRP3) related to PANoptosis in SCI; drug predictions may target hub genes for SCI treatment (e.g., EMRICASAN) |
| Machine Learning and Experiments Revealed Key Genes Related to PANoptosis Linked to Drug Prediction and Immune Landscape in Spinal Cord Injury (Li et al. -2025) | 38 SCI patients, 10 healthy controls, 10 trauma controls | AIS grades A, B, C, D (acute stage) | GEO dataset (GSE151371)<br>Rats qRT-PCR data | Artificial Neural Network (ANN)<br>Random Forest | Inflammatory signaling, pyroptosis, apoptosis, necroptosis | ZnO NPs protect neurons from apoptosis post SCI in animal models, improve mitochondrial function, and modulate oxidative stress; potential for therapeutic use in SCI. |
| Protective effect of zinc oxide nanoparticles on spinal cord injury (Liu et al. -2022) | 38 SCI blood samples, 10 control blood samples in humans<br>6 SCI + 6 sham (GSE93561); 8 SCI + | 0.7N force at T10 | Gene Expression Omnibus (GEO) - GSE151371 + Lab data from rats | Machine learning models (RF, LASSO, SVM) | PI3K/Akt signaling pathway (both human and animal data) | Identification of key genes (SLC31A1, DBT, DLST, LIAS) associated with cuproptosis and their role in SCI; machine learning models predict gene expression and immune response SLC31A1 had the |

| | | | | | | |
|---|---|---|---|---|---|---|
| | 2 controls (GSE45376); 3 SCI + 3 sham (qRT-PCR) in mice | | | | | highest diagnostic value |
| Machine learning-driven prediction model for cuproptosis-related genes in spinal cord injury: construction and experimental validation (Zhou et al. - 2025) | 48 (38 SCI patients, 10 healthy controls) Rats Not specified | AIS grades A–D in the GSE151371 dataset | GSE151371 dataset + Rat qRT-PCR | Machine learning models (LASSO regression, SVM, Random Forest) | Cuproptosis-related (TCA cycle, mitochondrial metabolism) and Ribosome, HIF-1 signaling pathway | MSC transplantation reduces inflammation in SCI by inhibiting lactylation-related DEGs, improving clinical relevance |
| Mesenchymal stem cell transplantation ameliorates inflammation in spinal cord injury by inhibiting lactylation-related genes (Zou et al., 2025) | The GSE226238 dataset has 37 SCI and 9 healthy controls. The GSE151371 dataset, 58 patients: 38 with SCI, 10 healthy controls, and 10 others | Not specified *check | GEO datasets GSE226238 and GSE151371 | | T cell receptor signaling, NF-kB signaling, Th1/Th2 cell differentiation, mTOR signaling | |
| | | Not specified *check* | | | | |

13.D. Synthesis Methods (Statistical Synthesis Methods): As the effects of functional appliance treatment were deemed to be highly variable according to the patient's age, sex, and individual maturation of spinal cord injury. We based our primary analyses upon consideration of dichotomous process adherence measures (for instance the proportion of patients managed according to evidence-based recommendations). In order to provide a quantitative assessment of the effects associated with reminders without resorting to numerous assumptions orconverying a misleading degree of confidence in the results. The study designs contain bioinformatics analysis, experimental validation, retrospective analysis of prospective cohort, cross-sectional with invivo/invitro follow up, transcriptome analysis, qPCR validation, immune profiling, registered clinical trial, biomarker validation,

13.E.Methods to explore heterogeneity:

13.F. Synthesis Methods (Sensitivity Analyses):

14. Reporting Bias Assessment:

15. Certainty Assessment:

16. A. Study Selection (Flow of studies):

16.B. Study Selection (Excluded studies):

17. Study Characteristics:

18. Risk of Bias in studies:

19. Results of Individual studies:

20. A. Results of syntheses (Characteristics of Contributing Studies):

20. B.Results of syntheses (Results of statistical syntheses):

20 .C.Results of syntheses (results of investigations of heterogeneity):

20. D.Results of Syntheses (Results of sensitivity analyses):

21. Reporting Biases:

22. Certainty of Evidence:

23. A.Discussion (Interpretation):

23. B. Discussion (Limitations of Evidence):

23. C. Discussion (Limitations of review processes):

23.D. Discussion (Implications):

24. A. Registration and Protocol(Registration):

24.B. Registration and protocol (protocol):

24. C. Registration and protocol (amendments):

25. Support:

26. Competing interests:

27. Availability of data, code, and other materials:
