## Supplementory for "Artificial Intelligence Models for Predicting Molecular Pathway Activity in Spinal Cord Injury: A Systematic Review"

1. Specify your systematic review question:

| Criteria | Specify Systematic Review questions |
|---|---|
| Intended use of the model | Target Population: Humans with SCI<br>Outcome Predicted: Prediction of molecular signaling pathways and repair in humans with spinal cord injury<br>Time Horizon: During hospital stay<br>Clinical purpose: Predictive applications of AI in SCI molecular signaling pathways, repair, and prognostic processes in human SCI treatment<br>Setting: Primary care, emergency setting, ICU |
| Participants, including selection criteria and setting: | Humans with SCI and AI prediction and signaling pathways:<br>(Spinal cord injury or SCI or spinal trauma or myelopathy) and ("artificial intelligence" OR "AI" OR "machine learning" OR "deep learning" OR "neural network" OR "predictive model" OR "random forest" OR "support vector machine" OR "SVM") and "molecular pathway" OR "signaling pathway" OR "gene network" OR "transcriptomic" OR "proteomic" OR "metabolomic" OR "bioinformatic" OR "systems biology" ) |
| Predictors (used in prediction modelling), including types of predictors (e.g. history, clinical examination, biochemical markers, imaging tests), time of measurement, specific measurement issues (e.g., any requirements/ prohibitions for specialized equipment): | Humans with spinal cord injury or spinal cord trauma, and stem cell interventions or molecular signaling pathways or proteomic or genomic or bioinformatics analysis, and artificial intelligence prediction or neural network in predictive usages<br>Time of measurement: after injury and before intervention or after injury and after intervention |
| Outcome to be predicted: | SCI repair, SCI treatment, molecular signaling pathways after SCI |

2.Classify the type of Prediction Model Evaluation:

| | | | |
|---|---|---|---|
| Classify the evaluation based on its aim | | | |
| Type of prediction study | PROBAST boxes to complete | Tick as appropriate | Definition for type of prediction model study |
| Development only | | | A study that creates a new prediction model from scratch using clinical data. |

| | | | |
|---|---|---|---|
| Development and validation | Yes, To investigate new prediction models | Yes | Develop a new prediction model from clinical data and then validate it either internally(same data sources) or externally (different data sources) |
| Validation only | | | A study that tests an existing model on a new patient population to assess performance. |

| Publication | Models of Interest | Outcomes of Interest |
|---|---|---|

Step 3. Assess risk of bias and applicability:

Study 1

### DOMAIN 1: Participants

**A. Risk of Bias**

- **Sources of data and criteria for participant selection:**
    - Development: TCGA gastric cancer cohort (RNA-seq + clinical survival data).
    - Validation: Multiple GEO cohorts (GSE datasets).
    - Inclusion: Patients with transcriptome data and overall survival.
    - Exclusion: Cases lacking survival or clinical information.

| Signalling question | Dev | Val |
|---|---|---|
| 1.1 Appropriate data sources used? | Y | Y |
| 1.2 All inclusions/exclusions appropriate? | Y | Y |

**Risk of bias: Low**
**Rationale:** Data came from large, high-quality public cohorts with transparent inclusion criteria.

**B. Applicability**

- **Participants, setting, dates:** Gastric cancer patients in TCGA and GEO (diagnoses ~2008–2019).
- **Concern:** Low
- **Rationale:** Patient population representative of gastric cancer prognosis studies.

---

### DOMAIN 2: Predictors

**A. Risk of Bias**

- **Predictors included in final model:** Immune-related gene pairs (IRGPs) constructed from transcriptomic expression of immune-related genes.
- **Definition/timing:** All predictors derived at baseline from tumor samples before the outcome is known.

| Signalling question | Dev | Val |
|---|---|---|
| 2.1 Predictors defined/assessed similarly for all? | Y | Y |
| 2.2 Predictor assessments blinded to outcome? | PY | PY |
| 2.3 Predictors available at intended time of use? | Y | Y |

**Risk of bias: Low/Unclear**
**Rationale:** Predictors were consistently defined and reproducible, but retrospective data mean blinding to outcomes cannot be guaranteed.

**B. Applicability**

- **Concern:** Low
- **Rationale:** Predictors (immune-related genes) are relevant and available in clinical-translational research settings.

### DOMAIN 3: Outcome

**A. Risk of Bias**

- **Outcome definition:** Overall survival (OS).
- **Determination:** Time from diagnosis to death or last follow-up.
- **Timing:** Longitudinal follow-up available in TCGA/GEO.

| Signalling question | Dev | Val |
|---|---|---|
| 3.1 Outcome determined appropriately? | Y | Y |
| 3.2 Pre-specified/standard definition used? | Y | Y |
| 3.3 Predictors excluded from outcome definition? | Y | Y |
| 3.4 Outcome defined similarly for all? | Y | Y |
| 3.5 Outcome determined without knowledge of predictors? | PY | PY |
| 3.6 Time interval appropriate? | Y | Y |

**Risk of bias: Low**
**Rationale:** OS is a robust, standardized, objective endpoint; the retrospective nature introduces only minor concern about blinding.

**B. Applicability**

- **Outcome timing:** Death/last follow-up; no composite endpoint.
- **Concern:** Low

- **Rationale:** OS is directly aligned with clinical prognosis assessment.

#### DOMAIN 4: Analysis

**Risk of Bias**

- **Participants/predictors:** Hundreds of patients in TCGA; multiple GEO external cohorts.
- **Development:** LASSO Cox regression on IRGPs; predictors reduced via penalization.
- **Validation:** External validation in independent GEO cohorts.
- **Performance measures:** ROC, Kaplan–Meier survival, C-index, calibration.
- **Missing data:** Not explicitly described.
- **Overfitting:** Addressed via LASSO and external validation.

| Signalling question | Dev | Val |
|---|---|---|
| 4.1 Reasonable number with outcome? | Y | Y |
| 4.2 Predictors handled appropriately? | Y | Y |
| 4.3 All enrolled participants included? | Y | Y |
| 4.4 Missing data handled appropriately? | NI | NI |
| 4.5 Predictor selection not solely univariable? | Y | Y |
| 4.6 Complexities (e.g. censoring) handled? | Y | Y |
| 4.7 Performance measures appropriate? | Y | Y |
| 4.8 Overfitting/optimism accounted for? | Y | Y |
| 4.9 Predictors' weights correspond to multivariable results? | Y | Y |

**Risk of bias: Low/Unclear**
**Rationale:** Strong methodological approach and robust external validation, but unclear reporting of missing data handling prevents a full low-risk rating.

Study 2

#### DOMAIN 1: Participants

##### A. Risk of Bias

- Data: TCGA-CRC cohort for development; GEO datasets for validation.
- Inclusion: Patients with transcriptomic and survival data.
- Exclusion: Patients lacking follow-up or clinical data.

| Question | Dev | Val |
|---|---|---|
| 1.1 Appropriate data sources? | Y | Y |
| 1.2 Inclusions/exclusions appropriate? | Y | Y |

**Risk of Bias:** Low
**Rationale:** Standardized high-quality cohorts; transparent criteria.

##### B. Applicability

- Participants: Colorectal cancer patients, various stages.
- Dates: TCGA (~2008–2018), GEO earlier cohorts.
  **Concern:** Low
  **Rationale:** Matches CRC prognostic population.

---

#### DOMAIN 2: Predictors

##### A. Risk of Bias

- Predictors: Immune-related genes (expression signatures).
- Timing: Baseline tumor data.
- Assessment: Consistent across cohorts.

| Question | Dev | Val |
|---|---|---|
| 2.1 Predictors assessed similarly? | Y | Y |
| 2.2 Predictor assessment blinded to outcome? | PY | PY |
| 2.3 Predictors available at intended use? | Y | Y |

**Risk of Bias:** Low/Unclear
**Rationale:** Retrospective data = possible lack of blinding.

**B. Applicability**
**Concern:** Low
**Rationale:** Predictors relevant to immunogenomic prognostics.

---

#### DOMAIN 3: Outcome

**A. Risk of Bias**

- Outcome: Overall survival (OS).
- Determination: Time from diagnosis → death or last follow-up.
- Standard across cohorts.

| Question | Dev | Val |
|---|---|---|
| 3.1 Outcome appropriate? | Y | Y |
| 3.2 Standard definition used? | Y | Y |
| 3.3 Predictors excluded from outcome definition? | Y | Y |
| 3.4 Outcome measured similarly? | Y | Y |
| 3.5 Outcome determined without predictor knowledge? | PY | PY |
| 3.6 Time interval appropriate? | Y | Y |

**Risk of Bias:** Low
**Rationale:** OS is objective, robust, and standardized.

**B. Applicability**

- Timepoint: OS only, no composite.

**Concern:** Low
**Rationale:** Fully aligns with prognostic question.

---

#### DOMAIN 4: Analysis

- N: Adequate sample size; hundreds of CRC cases.
- Model: LASSO Cox regression; penalization to reduce overfitting.
- Validation: External GEO cohorts.
- Performance: C-index, ROC, KM survival, calibration.
- Missing data: Not always explicitly reported.

| Question | Dev | Val |
|---|---|---|
| 4.1 Enough participants/outcomes? | Y | Y |
| 4.2 Predictors handled appropriately? | Y | Y |
| 4.3 All participants included? | Y | Y |
| 4.4 Missing data handled appropriately? | NI | NI |
| 4.5 Avoided univariable-only selection? | Y | Y |
| 4.6 Complexities handled? | Y | Y |
| 4.7 Performance measures adequate? | Y | Y |
| 4.8 Overfitting addressed? | Y | Y |
| 4.9 Final model weights from multivariable analysis? | Y | Y |

**Risk of Bias:** Low/Unclear
**Rationale:** Very good design, external validation, but the missing data handling is unclear.

---

#### Step 4: Overall Assessment (CRC model)

- **Risk of Bias:** Low/Unclear
- **Applicability Concern:** Low

- **Summary:** Model robust and externally validated, with minor uncertainties in retrospective data handling.

---

#### DOMAIN 1: Participants

##### A. Risk of Bias

- Data: TCGA-STAD for development; multiple GEO datasets for validation.
- Inclusion: Patients with RNA-seq + OS data.
- Exclusion: Missing survival info.

| Question | Dev | Val |
|---|---|---|
| 1.1 Appropriate sources? | Y | Y |
| 1.2 Inclusions/exclusions appropriate? | Y | Y |

**Risk of Bias:** Low

##### B. Applicability

- Gastric cancer patients, multi-cohort.
  **Concern:** Low

---

#### DOMAIN 2: Predictors

##### A. Risk of Bias

- Predictors: Immune-related gene pairs (IRGPs).
- Timing: Baseline tumor data.

| Question | Dev | Val |
|---|---|---|
| 2.1 Predictors defined | Y | Y |

| | | |
|---|---|---|
| consistently? | | |
| 2.2 Blinded to outcome? | PY | PY |
| 2.3 Available at intended use? | Y | Y |

**Risk of Bias:** Low/Unclear
**Rationale:** Retrospective = possible outcome knowledge bias.

**B. Applicability**
**Concern:** Low
**Rationale:** Predictors relevant to clinical-translational GC research.

---

#### DOMAIN 3: Outcome

**A. Risk of Bias**

- Outcome: OS.
- Determination: Diagnosis → death/last follow-up.

| Question | Dev | Val |
|---|---|---|
| 3.1 Outcome appropriate? | Y | Y |
| 3.2 Standard definition? | Y | Y |
| 3.3 Predictors excluded? | Y | Y |
| 3.4 Similar across participants? | Y | Y |
| 3.5 Outcome determined w/o predictor info? | PY | PY |
| 3.6 Interval appropriate? | Y | Y |

**Risk of Bias:** Low

**B. Applicability**
**Concern:** Low
**Rationale:** OS directly fits the prognostic review question.

---

#### DOMAIN 4: Analysis

- N: Hundreds of gastric cancer cases.

- Model: LASSO Cox regression.
- Validation: Multiple external GEO cohorts.
- Performance: ROC, KM survival, calibration, C-index.
- Missing data: Not fully reported.

| Question | Dev | Val |
|---|---|---|
| 4.1 Enough participants/outcomes? | Y | Y |
| 4.2 Predictors handled appropriately? | Y | Y |
| 4.3 All participants included? | Y | Y |
| 4.4 Missing data handled? | NI | NI |
| 4.5 Univariable-only selection avoided? | Y | Y |
| 4.6 Complexities handled? | Y | Y |
| 4.7 Performance measures appropriate? | Y | Y |
| 4.8 Overfitting addressed? | Y | Y |
| 4.9 Final weights match multivariable results? | Y | Y |

**Risk of Bias:** Low/Unclear

---

##### Step 4: Overall Assessment (GC model)

- **Risk of Bias:** Low/Unclear
- **Applicability Concern:** Low
- **Summary:** Well-developed, multi-cohort external validation. Uncertainty mainly in missing data handling.

Study 3

### DOMAIN 1: Participants

##### A. Risk of Bias

| Signalling Question | Answer | Notes |
|---|---|---|
| 1.1 Appropriate data sources (cohort, case-control, etc.)? | Y | Public GEO case-control transcriptomic datasets were used. |
| 1.2 Were inclusions/exclusions appropriate? | PY | Preprocessing described, but some filtering criteria not fully transparent. |

**Risk of bias rating: Low–Unclear**
**Rationale:** Use of standardized datasets is appropriate, but inclusion/exclusion methods only partially described.

##### B. Applicability

| Aspect | Concern | Rationale |
|---|---|---|
| Participants, setting, dates | Low–Unclear | Human NDD patients and controls are relevant, but multiple GEO cohorts introduce heterogeneity in setting/time. |

---

#### DOMAIN 2: Predictors

##### A. Risk of Bias

| Signalling Question | Answer | Notes |
|---|---|---|
| 2.1 Predictors defined/assessed similarly? | PY | Batch effects across datasets could cause inconsistencies. |
| 2.2 Predictor assessments blinded to outcomes? | Y | Gene expression measured independently of diagnosis. |
| 2.3 Predictors available at intended time of use? | Y | Transcriptomic data could be measured at baseline. |

**Risk of bias rating: Low–Unclear**
**Rationale:** Predictors valid, but cross-dataset variation may bias results.

##### B. Applicability

| Aspect | Concern | Rationale |
|---|---|---|
| Definition, assessment, timing of predictors | Low | Biomarkers chosen (hub genes, immune infiltration) are biologically relevant to NDD. |

---

### DOMAIN 3: Outcome

#### A. Risk of Bias

| Signalling Question | Answer | Notes |
| --- | --- | --- |
| 3.1 Was the outcome determined appropriately? | Y | Outcomes based on diagnostic status in original datasets. |
| 3.2 Pre-specified/standard outcome definition used? | PY | Diagnostic criteria varied across datasets. |
| 3.3 Were predictors excluded from outcome definition? | Y | No overlap with gene expression predictors. |
| 3.4 Outcome defined/determined similarly for all? | PY | Some variability in diagnostic annotation across studies. |
| 3.5 Outcome determined independently of predictors? | Y | Yes, diagnostic status was assigned separately. |
| 3.6 Time interval appropriate? | NI | Cross-sectional transcriptomic studies, no longitudinal follow-up. |

**Risk of bias rating: Unclear**
**Rationale:** Diagnosis validity differs across datasets, and no follow-up was considered.

#### B. Applicability

| Aspect | Concern | Rationale |
| --- | --- | --- |
| Outcome definition/timing | Low–Unclear | Outcomes relevant (NDD diagnosis), but heterogeneity across cohorts limits applicability. |

---

### DOMAIN 4: Analysis

#### A. Risk of Bias

| Signalling Question | Answer | Notes |
| --- | --- | --- |
| 4.1 Reasonable number of outcome events? | PY | Moderate sample sizes, some small subgroups. |
| 4.2 Predictors handled appropriately? | Y | Standard ML feature selection used. |
| 4.3 All participants included in analysis? | NI | Not fully reported. |

| | | |
|---|---|---|
| 4.4 Missing data handled appropriately? | NI | Missing data treatment not described. |
| 4.5 Univariable selection avoided? | Y | Multivariable ML algorithms applied. |
| 4.6 Data complexities handled (batch, censoring, etc.)? | PN | Batch effects not fully addressed. |
| 4.7 Model performance measured properly? | Y | AUC, ROC, calibration used. |
| 4.8 Overfitting/optimism adjusted for? | PN | No external validation, limited internal validation. |
| 4.9 Predictors consistent with multivariable results? | Y | Yes, hub genes confirmed across ML models. |

**Risk of bias rating: High**
**Rationale:** Lack of external validation, unclear missing data handling, batch effect risks, and possible overfitting.

---

### Step 4: Overall Assessment

| Category | Rating | Summary |
|---|---|---|
| **Overall Risk of Bias** | **High** | Mainly due to weaknesses in analysis (no external validation, batch effects, unclear missing data). |
| **Overall Applicability Concern** | **Low–Unclear** | Predictors and outcomes relevant to NDD, but dataset heterogeneity and diagnostic variability limit generalizability. |

Study 4

### Domain 1: Participants

| Section | Signaling Questions | Dev | Val |
|---|---|---|---|
| **Risk of Bias** | 1.1 Were appropriate data sources used? | Y | Y |
| | 1.2 Were all inclusions and exclusions appropriate? | Y | Y |
| **Risk of Bias Judgment** | | **Low** | **Low** |

| | | | |
|---|---|---|---|
| **Rationale** | Prospective multicenter cohort with clear inclusion/exclusion criteria, minimizing selection bias. | | |
| **Applicability** | Concern that included participants and setting do not match review question | **Low** | **Low** |
| **Rationale** | Participants (adults with neurological disorders) and setting (multicenter hospitals, 2022–2024) are appropriate for the review question. | | |

---

#### Domain 2: Predictors

| Section | Signaling Questions | Dev | Val |
|---|---|---|---|
| **Risk of Bias** | 2.1 Were predictors defined and assessed similarly for all participants? | Y | Y |
| | 2.2 Were predictor assessments made without knowledge of outcome data? | PY | PY |
| | 2.3 Are all predictors available at the time the model is intended to be used? | Y | Y |
| **Risk of Bias Judgment** | | **Low/Unclear** | **Low/Unclear** |
| **Rationale** | Predictors were standardized and clinically available, but some retrospective extraction raises minor concerns. | | |
| **Applicability** | Concern that definition/assessment/timing of predictors does not match review question | **Low** | **Low** |
| **Rationale** | Predictors (clinical, imaging, lab biomarkers) are consistent with clinical practice and relevant to intended use. | | |

---

#### Domain 3: Outcome

| Section | Signaling Questions | Dev | Val |
|---|---|---|---|
| **Risk of Bias** | 3.1 Was the outcome determined appropriately? | Y | Y |
| | 3.2 Was a pre-specified/standard definition used? | Y | Y |

| | | | |
|---|---|---|---|
| | 3.3 Were predictors excluded from outcome definition? | Y | Y |
| | 3.4 Was outcome defined/determined similarly for all participants? | Y | Y |
| | 3.5 Was outcome determined without knowledge of predictors? | PY | PY |
| | 3.6 Was time interval appropriate? | Y | Y |
| **Risk of Bias Judgment** | | **Low** | **Low** |
| **Rationale** | Outcomes (12-month neurological prognosis with validated scales) were clearly defined and consistently assessed. Blinding of assessors was only partial. | | |
| **Applicability** | Concern that outcome definition/timing does not match review question | **Low** | **Low** |
| **Rationale** | Outcome is clinically meaningful and measured at relevant follow-up time. | | |

---

#### Domain 4: Analysis

| Section | Signaling Questions | Dev | Val |
|---|---|---|---|
| **Risk of Bias** | 4.1 Were there enough participants with outcome? | Y | Y |
| | 4.2 Were continuous and categorical predictors handled appropriately? | Y | Y |
| | 4.3 Were all enrolled participants included? | Y | Y |
| | 4.4 Were participants with missing data handled appropriately? | Y | Y |
| | 4.5 Was univariable predictor selection avoided? | Y | Y |
| | 4.6 Were data complexities accounted for? | Y | Y |
| | 4.7 Were model performance measures evaluated appropriately? | Y | Y |
| | 4.8 Was overfitting/optimism accounted for? | Y | Y |
| | 4.9 Do predictors/weights correspond to multivariable results? | Y | Y |

| | | | |
|---|---|---|---|
| **Risk of Bias Judgment** | | **Low** | **Low** |
| **Rationale** | Large sample size, adequate events per predictor, appropriate modeling (LASSO logistic regression), internal validation with bootstrapping and temporal split, optimism-adjusted calibration/discrimination metrics, and missing data imputed. | | |

---

#### Step 4: Overall Assessment

| Category | Judgment | Rationale |
|---|---|---|
| **Overall Risk of Bias** | **Low** | All domains low risk, except minor uncertainty in blinding (predictors/outcomes). Strong internal validation reduces bias. |
| **Overall Concern for Applicability** | **Low** | Participants, predictors, and outcomes align with intended clinical use and review question. |
