## Supplementory for "Artificial Intelligence Models for Predicting Molecular Pathway Activity in Spinal Cord Injury: A Systematic Review"

1. Specify Your Systematic Review Question

| **Criteria** | Specify Your Systematic Review Question |
|---|---|
| **Intended use of model** | To identify high-confidence molecular pathways for:<br>- Mechanistic understanding of SCI progression.<br>- Biomarker discovery<br>- Therapeutic target prioritization |
| **Participants, including selection criteria and setting** | Inclusion Criteria:<br>- Human participants with SCI (any severity/phase) or human-derived omics data (e.g., from GEO/SRA databases).<br>- Studies using AI/ML to analyze molecular pathways.<br><br>Exclusion Criteria:<br>- Rodent-only studies.<br>- Non-AI approaches<br><br>Setting:<br>- Clinical cohorts or computational studies |
| **Predictors (used in prediction modeling), including types of predictors (e.g. history, clinical examination, biochemical markers, imaging tests), time of measurement, specific measurement issues (e.g., any requirements/ prohibitions for specialized equipment)** | Types of Predictors:<br>- Omics features: Gene expression (RNA-seq), protein levels (mass spectrometry), metabolites.<br>- Clinical/demographic variables (if available): Age, injury severity (ASIA score), time since injury.<br><br>Time of Measurement:<br>- Acute (<72h), subacute (1–4 weeks), or chronic (>1 month) post-SCI. |
| **Outcome to be predicted** | - Activation status of molecular pathways<br><br>- Pathway-associated clinical outcomes |

2.Classify the type of Prediction Model Evaluation:

| Classify the evaluation based on its aim | | | |
|---|---|---|---|
| **Type of prediction study** | **PROBAST boxes to complete** | **Tick as appropriate** | **Definition for type of prediction model study** |
| **Development only** | **Development** | | **A study that creates a new prediction model from scratch using clinical data.** |
| **Development and validation** | **Development and validation** | ✅ | **Develop a new prediction model from clinical data and then validate it either internally(same data sources) or externally (different datasources)** |
| **Validation only** | **Validation** | | **A study that tests an existing model on a new patient population to assess performance.** |

| | |
|---|---|
| **Publication reference** | Zhang, Z., Zhu, Z., Wang, X., Liu, D., Liu, X., Mi, Z., Tao, H., & Fan, H. (2023). Comprehensive landscape of immune-based classifier related to early diagnosis and macrophage M1 in spinal cord injury. AGING, 15(4), 1158–1176. https://doi.org/10.18632/aging.204548 |
| **Models of interest** | Immune-based classifier for early diagnosis of spinal cord injury (SCI), focusing on two core immune-related genes (IRGs): FCER1G and NFATC2, identified via machine learning (Random Forest, LASSO) and PPI analysis. |
| **Outcome of interest** | Early diagnosis of SCI and association with immune microenvironment dysregulation (e.g., macrophage M1 polarization). The model predicts SCI occurrence with high accuracy (AUC = 1.000 for combined IRGs). |

| Classify the evaluation based on its aim |
|---|

| Type of prediction study | PROBAST boxes to complete | Tick as appropriate | Definition for type of prediction model study |
|---|---|---|---|
| **Development only** | **Development** | | **A study that creates a new prediction model from scratch using clinical data.** |
| **Development and validation** | **Development and validation** | ✅ | **Develop a new prediction model from clinical data and then validate it either internally(same data sources) or externally (different datasources)** |
| **Validation only** | **Validation** | | **A study that tests an existing model on a new patient population to assess performance.** |

| | |
|---|---|
| **Publication reference** | Li J, Liu X, Wang J, et al. (2023). "Identification of immunodiagnostic blood biomarkers associated with spinal cord injury severity." Frontiers in Immunology, 14:1101564. DOI: 10.3389/fimmu.2023.1101564 |
| **Models of interest** | The study identifies immunodiagnostic blood biomarkers for spinal cord injury (SCI) and its severity grades (AIS A and AIS D) using weighted gene coexpression network analysis (WGCNA) and least absolute shrinkage and selection operator (LASSO) logistic regression. |
| **Outcome of interest** | The outcomes include the diagnostic value of identified biomarkers (e.g., CKLF, EDNRB, FCER1G, SORT1, TNFSF13B for SCI; GDF11, HSPA1L for AIS A; PRKCA, CMTM2 for AIS D) and their correlation with immune cell changes in blood post-SCI. The diagnostic sensitivity was evaluated using the |

| | |
|---|---|
| | area under the curve (AUC) of receiver operating characteristic (ROC) curves. |

3. Assess risk of bias and applicability:

Zhang, Z., 2023.

| DOMAIN 1: Participants | | | |
|---|---|---|---|
| **A. Risk of Bias** | | | |
| **Describe the sources of data and criteria for participant selection:**<br>Data sources: Microarray expression profile for SCI patients was acquired from GSE151371 in the GEO database. Peripheral blood samples from 10 SCI patients and 8 HC were collected from Xi-Jing Hospital<br>Criteria for participant selection: Not specified | | | |
| | | **Dev** | **Val** |
| **1.1 Were appropriate data sources used, e.g. cohort, RCT or nested case-control study data?** | | Y | Y |
| **1.2 Were all inclusions and exclusions of participants appropriate?** | | PY | PY |
| **Risk of bias introduced by selection of participants:** | **RISK:<br>(low/ high/ unclear)** | unclear | unclear |
| **Rationale of bias rating:**<br>Development: Reliance on public data (GSE151371) limits clarity on participant recruitment (e.g., severity, timing of sample collection post-SCI).<br>Validation: Small external cohort and lack of detailed exclusion criteria (e.g., comorbidities, medications) introduce uncertainty. | | | |
| **B. Applicability** | | | |
| **Describe included participants, setting and dates:**<br>Development:<br>Participants: 38 SCI patients and 10 healthy controls (peripheral blood leukocytes).<br>Setting: Retrospective analysis of GEO data; original study setting/sampling dates not specified.<br>Validation:<br>Participants: 10 SCI patients (demographics in Supplementary Table 1) and 8 healthy controls from Xi-Jing Hospital.<br>Setting: Clinical samples + rat model (T10 spinal cord clamping); dates not reported. | | | |

| DOMAIN 1: Participants | | | |
|---|---|---|---|
| **Concern that the included participants and setting do not match the review question** | **Concern: (low/ high/ unclear)** | Low | Low |
| **Rationale of applicability rating:**<br>Data sources (transcriptomic profiles, peripheral blood) align with the study's aim to identify immune-related biomarkers for SCI.<br>External validation (human cohort + animal model) supports generalizability, though human sample size is small. | | | |

| DOMAIN 2: Predictors | | | |
|---|---|---|---|
| **A. Risk of Bias** | | | |
| **List and describe predictors included in the final model, e.g. definition and timing of assessment:**<br>The model includes two core immune-related genes (IRGs):<br>1. FCER1G (Fc epsilon receptor Ig):<br>Definition: Encodes the Fc receptor γ-chain, involved in pro-inflammatory immune responses (e.g., macrophage M1 polarization).<br>Timing of assessment: Measured in peripheral blood leukocytes (transcriptomic data) post-SCI (exact timing post-injury not specified in GEO data).<br>2. NFATC2 (Nuclear factor of activated T cells 2):<br>Definition: Transcription factor regulating T-cell activation and cytokine production.<br>Timing of assessment: Same as FCER1G. | | | |
| | | Dev | Val |
| **2.1 Were predictors defined and assessed in a similar way for all participants?** | | Y | PY |
| **2.2 Were predictor assessments made without knowledge of outcome data?** | | NI | PN |
| **2.3 Are all predictors available at the time the model is intended to be used?** | | Y | Y |
| **Risk of bias introduced by predictors or their assessment** | **RISK: (low/ high/ unclear)** | Unclear | Unclear |
| **Rationale of bias rating:**<br>Development: Lack of blinding details in GEO data.<br>Validation: Potential technical variability in qPCR/WB/IF protocols (e.g., antibody specificity, | | | |

| DOMAIN 2: Predictors | | | |
|---|---|---|---|
| normalization methods). | | | |
| **B. Applicability** | | | |
| **Concern that the definition, assessment or timing of predictors in the model do not match the review question** | **Concern: (low/ high/ unclear)** | Low | Low |
| **Rationale of applicability rating:**<br>Predictors (FCER1G/NFATC2) are biologically plausible for immune response post-SCI and measurable in clinical labs.<br>Timing of assessment (post-injury) aligns with the goal of early diagnosis, though exact clinical window (e.g., acute vs. subacute phase) is unclear. | | | |

| DOMAIN 3: Outcome | | |
|---|---|---|
| **A. Risk of Bias** | | |
| **Describe the outcome, how it was defined and determined, and the time interval between predictor assessment and outcome determination:**<br>The study's primary outcome was the diagnosis of spinal cord injury (SCI) based on immune-related gene (IRG) expression patterns (FCER1G and NFATC2), validated through:<br>Development phase: Differential gene expression in SCI vs. healthy controls (GSE151371 dataset).<br>Validation phase:<br>qPCR in an external human cohort (10 SCI patients, 8 controls).<br>In vivo experiments (rat SCI model: WB, IF, sequencing at 3/7 days post-injury). | | |
| | **Dev** | **Val** |
| **3.1 Was the outcome determined appropriately?** | Y | PY |
| **3.2 Was a pre-specified or standard outcome definition used?** | Y | PN |
| **3.3 Were predictors excluded from the outcome definition?** | Y | Y |
| **3.4 Was the outcome defined and determined in a similar way for all** | Y | PY |

<table>
<tr><td colspan="4">DOMAIN 3: Outcome</td></tr>
<tr><td colspan="2">participants?</td><td></td><td></td></tr>
<tr><td colspan="2">3.5 Was the outcome determined without knowledge of predictor information?</td><td>NI</td><td>PN</td></tr>
<tr><td colspan="2">3.6 Was the time interval between predictor assessment and outcome determination appropriate?</td><td>NI</td><td>PY</td></tr>
<tr><td>Risk of bias introduced by the outcome or its determination</td><td>RISK:<br>(low/ high/ unclear)</td><td>Unclear</td><td>Unclear</td></tr>
<tr><td colspan="4">Rationale of bias rating:<br>Development: Lack of details on SCI severity/timing in GEO data.<br>Validation: No blinding in lab assays; human cohort lacks standardized diagnostic timing.</td></tr>
<tr><td colspan="4">B. Applicability</td></tr>
<tr><td colspan="4">At what time point was the outcome determined:<br>Development: Unclear (retrospective GEO data).<br>Validation: Rats at 3/7 days post-injury; humans at unspecified time post-SCI.<br><br>If a composite outcome was used, describe the relative frequency/distribution of each contributing outcome:<br>Not applicable</td></tr>
<tr><td>Concern that the outcome, its definition, timing or determination do not match the review question</td><td>CONCERN: (low/ high/ unclear)</td><td>Low</td><td>Low</td></tr>
<tr><td colspan="4">Rationale of applicability rating:<br>Outcome (SCI diagnosis) aligns with the review question of early immune-based detection.<br>Limitations: Human cohort lacks clinical detail (e.g., injury severity/phase), but rat model supports biological plausibility.</td></tr>
</table>

| DOMAIN 4: Analysis |
|---|
| **A. Risk of Bias** |
| **Describe numbers of participants, number of candidate predictors, outcome events and events per candidate predictor:** |

DOMAIN 4: Analysis

Development: 38 SCI patients vs. 10 healthy controls (GSE151371 dataset).
Validation:
Human: 10 SCI patients vs. 8 controls (external cohort).
Animal: Rat SCI model (sample size not specified).
Candidate predictors: 51 immune-related genes (IRGs) initially screened; final model = 2 predictors (FCER1G, NFATC2).
Outcome events: SCI diagnosis (binary: present/absent).
Events per predictor: ~19:1 (38 SCI cases / 2 predictors), below the typical 10:1 rule but mitigated by machine learning techniques.

**Describe how the model was developed (for example in regards to modelling technique (e.g. survival or logistic modelling), predictor selection, and risk group definition):**
Technique:
Machine learning: Random Forest + LASSO regression for predictor selection.
PPI network analysis (Cytoscape) to identify hub genes.
Predictor selection:
Initial 2067 DEGs → 51 IRGs via WGCNA/ImmPort → 2 core IRGs (intersection of Random Forest, LASSO, PPI).
Risk group definition: Unsupervised clustering (k=2) based on IRG expression patterns (immunoreactive vs. immunosuppressive).

**Describe whether and how the model was validated, either internally (e.g. bootstrapping, cross validation, random split sample) or externally (e.g. temporal validation, geographical validation, different setting, different type of participants):**
Internal validation: Not mentioned (no bootstrapping/cross-validation).
External validation:
Independent human cohort: qPCR validation of FCER1G/NFATC2.
Animal model: WB/IF/sequencing in rats.

**Describe the performance measures of the model, e.g. (re)calibration, discrimination, (re)classification, net benefit, and whether they were adjusted for optimism:**
Discrimination: AUC = 1.000 (combined IRGs), 0.982 (FCER1G), 1.000 (NFATC2).
Calibration/optimism adjustment: Not reported.
Net benefit/reclassification: Not assessed.

**Describe any participants who were excluded from the analysis:**
NI (no explicit exclusions reported).

**Describe missing data on predictors and outcomes as well as methods used for missing data:**
Missing data: NI (assumed complete data for GEO samples; validation cohort not described).
Methods for missing data: NI

| | **Dev** | **Val** |
|---|---|---|
| **4.1 Were there a reasonable number of participants with the outcome?** | PN | PN |
| **4.2 Were continuous and categorical predictors handled** | Y | Y |

| DOMAIN 4: Analysis | | | |
|---|---|---|---|
| **appropriately?** | | | |
| **4.3 Were all enrolled participants included in the analysis?** | | NI | NI |
| **4.4 Were participants with missing data handled appropriately?** | | NI | NI |
| **4.5 Was selection of predictors based on univariable analysis avoided?** | | Y | |
| **4.6 Were complexities in the data (e.g. censoring, competing risks, sampling of controls) accounted for appropriately?** | | N | N |
| **4.7 Were relevant model performance measures evaluated appropriately?** | | PY | PN |
| **4.8 Were model overfitting and optimism in model performance accounted for?** | | PN | |
| **4.9 Do predictors and their assigned weights in the final model correspond to the results from multivariable analysis?** | | Y | |
| **Risk of bias introduced by the analysis** | **RISK:**<br>**(low/ high/ unclear)** | High | High |
| **Rationale of bias rating:**<br>Small sample size relative to predictors.<br>No internal validation or overfitting mitigation.<br>Lack of missing data handling. | | | |

Li J, 2023

| DOMAIN 1: Participants | | |
|---|---|---|
| **A. Risk of Bias** | | |
| **Describe the sources of data and criteria for participant selection:**<br>Data sources: Microarray expression profile for SCI patients was acquired from GSE151371 in the GEO database. Blood samples were collected from 10 patients with SCI, 3 patients with closed noncentral nervous system trauma, and 3 healthy individuals from Seventh Affiliated Hospital of Sun Yat-sen University.<br>Criteria for participant selection: Not specified | | |
| | **Dev** | **Val** |

<table>
<tr><td colspan="2">1.1 Were appropriate data sources used, e.g. cohort, RCT or nested case-control study data?</td><td>Y</td><td>PY</td></tr>
<tr><td colspan="2">1.2 Were all inclusions and exclusions of participants appropriate?</td><td>PY</td><td>PN</td></tr>
<tr><td>Risk of bias introduced by selection of participants:</td><td>RISK:<br><br>(low/ high/ unclear)</td><td>unclear</td><td>High</td></tr>
<tr><td colspan="4">Rationale of bias rating:<br><br>Development: While the dataset is well-defined, potential biases (e.g., selection of SCI patients, timing of blood sampling) are not explicitly addressed.<br><br>Validation: Small validation cohort and unclear selection criteria increase bias risk.</td></tr>
<tr><td colspan="4">B. Applicability</td></tr>
<tr><td colspan="4">Describe included participants, setting and dates:<br>Development:<br>Participants: 38 SCI patients (12 AIS A, 4 AIS B, 6 AIS C, 11 AIS D) and 10 healthy controls and 10 Non-CNS Trauma controls.<br>Setting: Blood samples collected within ~30 hours post-injury.<br>Dates: Dataset retrieved from GEO; original collection dates not specified.<br>Validation:<br>Participants: 10 SCI, 3 TC, 3 HC (no AIS stratification provided).<br>Setting: Blood samples from Seventh Affiliated Hospital of Sun Yat-sen University. Dates not specified.</td></tr>
<tr><td>Concern that the included participants and setting do not match the review question</td><td>Concern:<br><br>(low/ high/ unclear)</td><td>Low</td><td>High</td></tr>
<tr><td colspan="4">Rationale of applicability rating:<br><br>Development:The dataset includes relevant SCI severity groups and controls, but the lack of detailed clinical characteristics (e.g., injury mechanism, comorbidities) may limit generalizability.</td></tr>
</table>

| |
|---|
| Validation:The small, poorly characterized validation cohort limits confidence in generalizability. |

| DOMAIN 2: Predictors |
|---|
| **A. Risk of Bias** |
| **List and describe predictors included in the final model, e.g. definition and timing of assessment:**<br>The study identified two tiers of biomarkers for SCI diagnosis and severity grading:<br><br>A. General SCI Diagnosis (All Severities)<br>1. CKLF (Chemokine-like factor 1)<br>Definition: Pro-inflammatory chemokine linked to neutrophil recruitment.<br>Timing: Measured in blood ~30.3 ± 18.9 hours post-injury.<br>Role: Upregulated in SCI; correlates with neutrophil infiltration.<br><br>2. EDNRB (Endothelin receptor type B)<br>Definition: Receptor involved in vascular and immune responses.<br>Timing: Same as above.<br>Role: Associated with neutrophil activation and blood-spinal cord barrier disruption.<br><br>3. FCER1G (Fc epsilon receptor Ig)<br>Definition: Immune complex receptor on macrophages/neutrophils.<br>Timing: Same as above.<br>Role: Tied to innate immune activation post-SCI.<br><br>4. SORT1 (Sortilin 1)<br>Definition: Lysosomal sorting protein with neuroinflammatory roles.<br>Timing: Same as above.<br>Role: Linked to cytokine release and axonal degeneration.<br><br>5. TNFSF13B (TNF superfamily member 13B)<br>Definition: Cytokine regulating B-cell survival.<br>Timing: Same as above.<br>Role: Implicated in adaptive immune response post-SCI.<br><br>B. SCI Severity-Specific Biomarkers<br>1. AIS A (Most Severe):<br>GDF11 (Growth differentiation factor 11): Downregulated; anti-inflammatory role.<br>HSPA1L (Heat shock protein family A member 1 like): Upregulated; stress response marker.<br><br>2. AIS D (Mildest):<br>PRKCA (Protein kinase C alpha): Downregulated; cell survival signaling.<br>CMTM2 (CKLF-like MARVEL transmembrane domain-containing 2): Upregulated; immune cell adhesion. |

| DOMAIN 2: Predictors | | | |
|---|---|---|---|
| | | Dev | Val |
| **2.1 Were predictors defined and assessed in a similar way for all participants?** | | Y | PY |
| **2.2 Were predictor assessments made without knowledge of outcome data?** | | PY | NI |
| **2.3 Are all predictors available at the time the model is intended to be used?** | | Y | Y |
| **Risk of bias introduced by predictors or their assessment** | **RISK: (low/ high/ unclear)** | Low | Unclear |
| **Rationale of bias rating:**<br>Development:Standardized transcriptomic methods reduce variability, but lack of blinding confirmation introduces minor uncertainty.<br><br>Validation: Unclear blinding and small validation cohort increase uncertainty. | | | |
| **B. Applicability** | | | |
| **Concern that the definition, assessment or timing of predictors in the model do not match the review question** | **Concern: (low/ high/ unclear)** | Low | High |
| **Rationale of applicability rating:**<br><br>Development: Blood-based biomarkers are clinically feasible, but the timing of sampling (~30 hours post-injury) may limit early diagnostic utility.<br><br>Validation: The validation cohort's limited size and lack of AIS stratification reduce confidence in generalizability. | | | |

| DOMAIN 3: Outcome |
|---|
| **A. Risk of Bias** |

| | | | |
|---|---|---|---|
| **Describe the outcome, how it was defined and determined, and the time interval between predictor assessment and outcome determination:**<br><br>Primary Outcome: Spinal Cord Injury (SCI) severity, classified using the American Spinal Injury Association Impairment Scale (AIS) grades (AIS A (Complete injury) to D (Incomplete injury varying degrees of preserved motor/sensory function) in this study).<br><br>How the Outcome Was Determined: Clinical assessment by trained specialists.<br><br>Time Interval Between Predictor Assessment and Outcome Determination:<br><br>Development: Blood sampling for biomarkers: ~30.3 ± 18.9 hours post-injury.<br><br>AIS grading: Presumed concurrent with biomarker sampling (no explicit delay reported).<br><br>Validation: Unclear: No details on whether AIS grading was re-assessed at time of qPCR validation. | | | |
| | | **Dev** | **Val** |
| **3.1 Was the outcome determined appropriately?** | | Y | PN |
| **3.2 Was a pre-specified or standard outcome definition used?** | | Y | NI |
| **3.3 Were predictors excluded from the outcome definition?** | | Y | NI |
| **3.4 Was the outcome defined and determined in a similar way for all participants?** | | PY | NI |
| **3.5 Was the outcome determined without knowledge of predictor information?** | | NI | NI |
| **3.6 Was the time interval between predictor assessment and outcome determination appropriate?** | | Y | NI |
| **Risk of bias introduced by the outcome or its determination** | **RISK:**<br><br>**(low/ high/ unclear)** | Low | High |

| |
|---|
| **Rationale of bias rating:**<br><br>Development: Standard AIS grading reduces bias, but lack of blinding details introduces minor uncertainty.<br><br>Validation: Poor reporting on outcome definition and assessment in validation. |
| **B. Applicability** |
| **At what time point was the outcome determined:**<br><br>Development: Acute phase: AIS grading performed within hours to days post-injury (exact timing not specified).<br><br>Validation: Not reported for validation cohort.<br><br>**If a composite outcome was used, describe the relative frequency/distribution of each contributing outcome:**<br>Not applicable. |

| | | | |
|---|---|---|---|
| **Concern that the outcome, its definition, timing or determination do not match the review question** | **CONCERN: (low/ high/ unclear)** | Low | High |

| |
|---|
| **Rationale of applicability rating:**<br><br>Development: AIS is clinically relevant, but early grading (<72h) may not reflect long-term recovery.<br><br>Validation: Validation lacks clarity on outcome alignment with clinical AIS grades. |

| DOMAIN 4: Analysis |
|---|
| **A. Risk of Bias** |
| **Describe numbers of participants, number of candidate predictors, outcome events and events per candidate predictor:** |

| |
|---|
| Development:<br><br>Participants: 58 total (10 HC, 10 TC, 38 SCI [12 AIS A, 4 AIS B, 6 AIS C, 11 AIS D]).<br><br>Candidate predictors: 237 immune-related genes (DEIGs) initially, narrowed to 17 hub genes, then 5 final biomarkers (CKLF, EDNRB, FCER1G, SORT1, TNFSF13B).<br><br>Outcome events: SCI severity (AIS grades). Events per predictor: ~7.6 (38 SCI/5 predictors).<br><br>Validation:<br><br>Participants: 16 total (3 HC, 3 TC, 10 SCI [AIS grades unspecified]).<br><br>Predictors: Same 5 biomarkers validated via qPCR. |
| **Describe how the model was developed (for example in regards to modelling technique (e.g. survival or logistic modelling), predictor selection, and risk group definition):**<br><br>Technique:<br><br>- WGCNA: Identified co-expression modules correlated with SCI severity.<br><br>- LASSO regression: Selected top immune-related genes with diagnostic value.<br><br>- Predictor selection: Genes from the MEBrown module (highest SCI correlation) were prioritized.<br><br>- Risk group definition: Biomarkers stratified by AIS grades (AIS A/D highlighted). |
| **Describe whether and how the model was validated, either internally (e.g. bootstrapping, cross validation, random split sample) or externally (e.g. temporal validation, geographical validation, different setting, different type of participants):**<br><br>Internal: None (no bootstrapping/cross-validation reported).<br><br>External: Independent qPCR cohort (n=16), but underpowered and poorly characterized. |
| **Describe the performance measures of the model, e.g. (re)calibration, discrimination, (re)classification, net benefit, and whether they were adjusted for optimism:** |

Discrimination: AUCs for biomarkers (0.90–1.00 in development; qPCR validation showed significant expression differences).

Calibration/optimism adjustment: Not reported.

**Describe any participants who were excluded from the analysis:**

None reported.

**Describe missing data on predictors and outcomes as well as methods used for missing data:**

Not mentioned; assumed complete for transcriptomic data.

| | **Dev** | **Val** |
|---|---|---|
| **4.1 Were there a reasonable number of participants with the outcome?** | Y | PN |
| **4.2 Were continuous and categorical predictors handled appropriately?** | Y | Y |
| **4.3 Were all enrolled participants included in the analysis?** | Y | NI |
| **4.4 Were participants with missing data handled appropriately?** | NI | NI |
| **4.5 Was selection of predictors based on univariable analysis avoided?** | Y | |
| **4.6 Were complexities in the data (e.g. censoring, competing risks, sampling of controls) accounted for appropriately?** | Y | NI |
| **4.7 Were relevant model performance measures evaluated appropriately?** | PY | PN |

| | | | |
|---|---|---|---|
| **4.8 Were model overfitting and optimism in model performance accounted for?** | | PN | |
| **4.9 Do predictors and their assigned weights in the final model correspond to the results from multivariable analysis?** | | Y | |
| **Risk of bias introduced by the analysis** | **RISK:**<br><br>**(low/ high/ unclear)** | Unclear | High |
| **Rationale of bias rating:**<br><br>Development: Clinically relevant predictors, but optimism unaddressed.<br><br>Validation: Poor generalizability due to limited validation. | | | |

4: Overall Assessment

Zhang, Z., 2023.

| **Overall judgement about risk of bias and applicability of the prediction model evaluation** | | |
|---|---|---|
| **Overall judgement of risk of bias** | **RISK:**<br>**(low/ high/ unclear)** | High |
| **Summary of sources of potential bias:**<br>1. Participant Selection:<br>Development: Public GEO data lacked details on SCI severity/timing; small control group (n=10).<br>Validation: Small external cohort (n=10 SCI) with unclear exclusion criteria.<br><br>2. Predictor Assessment:<br>No blinding reported in validation lab assays (qPCR/WB/IF).<br>Unclear timing of predictor measurement post-SCI in humans.<br><br>3. Outcome Determination:<br>No clinical diagnostic standards (e.g., ASIA scale) in validation cohort.<br>Unclear if outcome assessors were blinded to predictor data.<br>4. Analysis:<br>Overfitting: No internal validation (e.g., bootstrapping) despite high-dimensional data (51 IRGs → 2 predictors).<br>Sample Size: Low events-per-predictor ratio (38 SCI cases / 51 initial predictors).<br>Missing Data: Handling not described. | | |

| Overall judgement about risk of bias and applicability of the prediction model evaluation | | |
|---|---|---|
| **Overall judgement of applicability** | **CONCERN:**<br>**(low/ high/ unclear)** | Low |
| **Summary of applicability concerns:**<br>1. Alignment with Review Question:<br>Predictors (FCER1G/NFATC2) are biologically plausible for immune response post-SCI and measurable in clinical settings.<br>Outcome (SCI diagnosis) matches the goal of early detection.<br><br>2. Limitations:<br>Clinical Translation: Unclear how transcriptomic biomarkers would integrate into current diagnostic workflows.<br>Timing: Lack of detail on optimal post-injury window for predictor assessment.<br>Population: GEO cohort may not reflect general SCI populations (e.g., heterogeneity in injury mechanisms). | | |

Li J, 2023

| Overall judgement about risk of bias and applicability of the prediction model evaluation | | |
|---|---|---|
| **Overall judgement of risk of bias** | **RISK:**<br>**(low/ high/ unclear)** | High |
| **Summary of sources of potential bias:**<br>Development Phase:<br>- No internal validation (e.g., bootstrapping) to address overfitting.<br>- Unclear blinding of outcome assessors to biomarker data.<br>- Missing data handling not reported.<br><br>Validation Phase:<br>- Extremely small sample (n=16) with poorly characterized outcomes.<br>- No recalibration of performance metrics (e.g., AUCs).<br>- Lack of transparency in AIS grading and timing. | | |
| **Overall judgement of applicability** | **CONCERN:**<br>**(low/ high/ unclear)** | High |
| **Summary of applicability concerns:**<br>1. Development Phase:<br>- Biomarkers sampled at ~30 hours post-injury; utility in hyper-acute (<6h) settings unknown.<br><br>2. Validation Phase:<br>- Validation cohort mismatched (no AIS stratification, small n=16).<br>- No evidence of generalizability to diverse populations or settings. | | |
