## Supplementory for "Artificial Intelligence Models for Predicting Molecular Pathway Activity in Spinal Cord Injury: A Systematic Review"

Table 1. summary of data sources, AI models, pathways, and key metrics across included SCI–AI studies

| Study (Year) | Country / Data Source | N (SCI / Control) | AI Method(s) | Main Molecular Targets | Principal Pathways / Mechanisms | Performance Metrics | Validation Type | Notable Limitations |
| --- | --- | --- | --- | --- | --- | --- | --- | --- |
| Zhang et al. 2023 (7) | China / GSE151371 | 38 / 10 | Bioinf + ML | FCER1G, NFATC2 | Immune activation (macrophage-related) | AUC = 1.000 | Internal | Small n; no prognostic data |
| Li et al. 2023 (8) | China / GSE151371 | Not reported | Bioinf + ML | AIS stage genes: CKLF, EDNRB, etc. | NF‑κB, VEGF, JAK‑STAT | AUC = 0.825–1.000 | qPCR (n=10 SCI) | No mechanistic validation; acute-phase focus |
| Kyritsis et al. 2021 (9) | USA / WBC RNA | 58 / Not applicable | ML diag model | RNA panels | Not applicable | Acc = 72.7%; AUC 0.865–0.938 | Internal | Few AIS B/C; acute only |
| Li et al. 2024 (10) | China / GSE151371+valid | 6 / 6 | ML | ANO10, BST1, ZFP36L2 | Not applicable | AUC = 0.793–0.870 | Internal | No post-mortem data; no luciferase assay |
| Zhou et al. 2025 (11) | China / GSE151371 + rats | Not reported | ML | SLC31A1, DBT, DLST, LIAS | Cuproptosis, mitochondrial dysfunction | AUC = 0.958 | Rat qPCR | Limited human validation; small sample size |
| Liu et al. 2022 (12) | China / GSE151371 + in vivo | Not reported | ANN | 10‑gene panel | Mitochondrial function | AUC = 0.974 | Rat | Small sample size; in vitro lacks systemic context |
| Zou et al. 2025 (13) | China / GSE151371 + rat | Not reported | ML | 7 lactylation genes | Epigenetic immune regulation | Not applicable | MSC therapy in rats | Not reported |
| Li et al. 2022 (14) | China / GSE151371 | Not reported | ML | CCR7 | Immune suppression | Not applicable | Not reported | Mechanism only; no therapy |
| Li et al. 2025 (15) | China / GSE151371 | Not reported | ML | CASP4, NLRP3 | PANoptosis | Not applicable | In silico drug repurposing | No experimental human data |
| Su et al. 2025 (16) | China / GSE151371 + rat | Not reported | ML | PINK1, SQSTM1 | Autophagy dysregulation | Not applicable | Docking & rat | Limited target validation |
| Zhang et al. 2025 (17) | China / GSE151371 | Not reported | ML | S100A8/A12, IL2RB | Chronic pain via immune infiltration | Not applicable | Not reported | Observational only |

Table 2. Diagnostic & Severity Classification Outcomes

| Study | Data Source / Sample | Methods | Key Biomarkers | Performance | Notes / Limitations |
| --- | --- | --- | --- | --- | --- |
| Zhang et al. (2023) | GSE151371 (38 SCI, 10 controls) | Bioinformatics + ML | FCER1G, NFATC2 | AUC = 1.000 | - Small sample size.  - Lack of clinical/prognostic data.  - Mechanistic role of FCER1G/ NFATC2 unclear. |
| Li et al. (2023) | GSE151371 | Bioinformatics + ML feature selection | SCI: CKLF, EDNRB, FCER1G, SORT1, TNFSF13B; AIS A: GDF11, HSPA1L, TNFRSF25; AIS D: PRKCA, CMTM2 | AUC = 0.825–1.000 | -Subgroup classification (AIS A vs D)  -Small qPCR cohort (n=10 SCI)  -No mechanistic validation;  -Acute-phase focus |
| Kyritsis et al. (2021) | WBC RNA (human, n=58) | Diagnostic Modeling (ML) | RNA profile panels | Accuracy 72.7%; AUC = 0.865 (AIS A), 0.938 (AIS D) | - Small sample size for AIS B/C (n = 4/6).  - No longitudinal outcome prediction (only acute phase).  - Potential batch effects in RNA-seq (addressed with ComBat). |
| Li et al. (2024) | GSE151371 + validation | ML models | ANO10, BST1, ZFP36L2 | AUC = 0.793–0.870 | - Small sample size for RNA-seq (6 SCI vs. 6 controls).  - No dual luciferase validation for miRNA-mRNA interactions.  - Lack of post-mortem spinal cord tissue data. |
| Zhou et al. (2025) | GSE151371 + rats | ML model | Cuproptosis-related: SLC31A1, DBT, DLST, LIAS | AUC = 0.958 | -Prognostic value suggested  - Small sample size (human and rat data).  - No control for rehabilitation or comorbidities in human data.  - Limited validation beyond qRT-PCR in rats. |
| Liu et al. (2022) | GSE151371 + in vivo | ANN | 10-gene panel | AUC = 0.974 | -Added nanoparticle validation  -Small sample size;  -Rat: In vitro model lacks systemic complexity |

| Study | Focus | Findings | Pathways / Mechanisms | Notes |
| --- | --- | --- | --- | --- |
| Li et al. (2023) | Immune dysregulation | Biomarker sets | NF-κB, VEGF, JAK-STAT, TLR signaling | Consistent immune infiltration |
| Zou et al. (2025) | Epigenetics | 7 lactylation-related genes | Immune cell infiltration, inflammation | Novel angle (lactylation) |
| Li et al. (2022) | Immune suppression | CCR7 downregulation | Tfh cell function, infection risk | Connects SCI with immune deficiency |
| Li et al. (2025) | Cell death | PANoptosis genes (CASP4, NLRP3) | PANoptosis, inflammasome | Identified targets for therapy |
| Zhou et al. (2025) | Cell death & metabolism | Cuproptosis-related genes | Mitochondrial dysfunction | Mechanistic + therapeutic overlap |
| Su et al. (2025) | Autophagy | PINK1/SQSTM1 imbalance | Dysregulated autophagic flux | Linked to neuronal death |
| Zhang et al. (2025) | Chronic outcomes | S100A8/A12, IL2RB | Immune infiltration → neuropathic pain | Links acute to chronic phase |

Table 4. Therapeutic Prediction & Experimental Validation

| Study | Strategy | Intervention / Drug | Validation | Outcome |
| --- | --- | --- | --- | --- |
| Li et al. (2025) | Drug repurposing | Emricasan (CASP4 inhibitor), Alaproclate (NLRP3 modulator) | Computational | Predicted efficacy in reducing inflammation |
| Su et al. (2025) | Drug repurposing | Imatinib binding PINK1 (ΔG = −10.9 kcal/mol) | In silico + rat SCI model | Improved autophagy flux |
| Liu et al. (2022) | Nanoparticle therapy | ZnO nanoparticles | Rat SCI model | ↓ apoptosis, ↑ mitochondrial function |
| Zhou et al. (2025) | Cuproptosis-targeted therapy | Mitochondrial protective agents | Rat SCI model | Mitigated mitochondrial damage |
| Zou et al. (2025) | Cellular therapy | Mesenchymal stem cell transplantation | Rat SCI model | ↓ inflammation, improved outcomes |

Table 5. The GRADE Assessment

| Certainty Category | n / 11 Studies | Representative Limitations |
| --- | --- | --- |
| ✦✧✧✧ Very Low | 8 | Very serious RoB; indirectness due to animal‑only validation; high AUC inflation risk in small cohorts |
| ✦✦✧✧ Low | 3 | Serious RoB; acute‑only phase; limited sample diversity |

.
