## Supplementory for "Artificial Intelligence Models for Predicting Molecular Pathway Activity in Spinal Cord Injury: A Systematic Review"

**Authors information:**

**Mahdi Mehmandoost**

Affiliation: Student Research Committee, School of Medicine, Shahid Beheshti University of Medical Sciences, Tehran, Iran

**Mahtab Jabbar**

Affiliation: Student Research Committee, School of Medicine, Tehran University of Medical Sciences, Tehran,

Iran.

**Behnaz Rahatijafarabadi**

Affiliation: Student Research Committee, School of Medicine, Golestan University of Medical

Sciences, Gorgan, Iran

**Mandana Mehrdad**

Affiliation: Isfahan University, The School of Advanced Medical Technologies, Image Processing Department

**Roozbeh Tavanaei**

Affiliation: Functional Neurosurgery Research Center, Shohada Tajrish Comprehensive Neurosurgical Center of Excellence, Shahid Beheshti University of Medical Sciences, Tehran, Iran

**Ibrahim Mohammadzadeh**

Affiliation: Functional Neurosurgery Research Center, Shohada Tajrish Comprehensive Neurosurgical Center of Excellence, Shahid Beheshti University of Medical Sciences, Tehran, Iran

**Sayeh Oveisi**

Affiliation: Functional Neurosurgery Research Center, Shohada Tajrish Comprehensive Neurosurgical Center of Excellence, Shahid Beheshti University of Medical Sciences, Tehran, Iran

**Alireza Zali, MD**

Affiliation: Functional Neurosurgery Research Center, Shohada Tajrish Comprehensive Neurosurgical Center of Excellence, Shahid Beheshti University of Medical Sciences, Tehran, Iran

**Saeed Oraee Yazdani, MD**

Affiliation: Functional Neurosurgery Research Center, Shohada Tajrish Comprehensive Neurosurgical Center of Excellence, Shahid Beheshti University of Medical Sciences, Tehran, Iran

**Farzan Fahim, MD**

Affiliation: Functional Neurosurgery Research Center, Shohada Tajrish Comprehensive Neurosurgical Center of Excellence, Shahid Beheshti University of Medical Sciences, Tehran, Iran

***Corresponding author**

Farzan Fahim, MD

Neurosurgery resident, Functional Neurosurgery Research Center, Shohada-E-Tajrish

Comprehensive Neurosurgical Center of Excellence, Shahid Beheshti University of Medical

Sciences, Tehran, Iran

**Running Title:** AI for Molecular Pathway in Spinal Cord Injury
